# Somatic and Stem Cell Bank to Study the Contribution of African Ancestry to Dementia: African iPSC Initiative

**DOI:** 10.1101/2025.01.24.25320911

**Authors:** Mahmoud B. Maina, Murtala B. Isah, Jacob A. Marsh, Zaid Muhammad, Larema Babazau, Abdulrahman Alkhamis Idris, Ekaterina Aladyeva, Nadia Miller, Emma Starr, Katherine J. Miller, Scott Lee, Miguel Minaya, Selina Wray, Oscar Harari, Baba W. Goni, Louise C. Serpell, Celeste M. Karch

## Abstract

**INTRODUCTION:** Africa, home to 1.4 billion people and the highest genetic diversity globally, harbors unique genetic variants crucial for understanding complex diseases like neurodegenerative disorders. However, African populations remain underrepresented in induced pluripotent stem cell (iPSC) collections, limiting the exploration of population-specific disease mechanisms and therapeutic discoveries.

**METHODS:** To address this gap, we established an open-access African Somatic and Stem Cell Bank.

**RESULTS:** In this initial phase, we generated 10 rigorously characterized iPSC lines from fibroblasts representing five Nigerian ethnic groups and both sexes. These lines underwent extensive profiling for pluripotency, genetic stability, differentiation potential, and Alzheimer’s disease and Parkinson’s disease risk variants. CRISPR/Cas9 technology was used to introduce frontotemporal dementia-associated *MAPT* mutations (P301L and R406W).

**DISCUSSION:** This collection offers a renewable, genetically diverse resource to investigate disease pathogenicity in African populations, facilitating breakthroughs in neurodegenerative research, drug discovery, and regenerative medicine.

## 1. Background

Africa, home to approximately 1.4 billion people and over 2,000 spoken languages (30% of the world’s living languages) [1], harbors the highest genetic diversity of all continents [2]. This genetic diversity has profound implications for understanding the genetic basis of complex diseases, including neurodegenerative disorders [3]. Genetic studies reveal significant population-specific differences in the frequency and distribution of disease-associated variants for neurodegenerative diseases (e.g. *APOE* and *ABCA7*) [4]. These genetic differences contribute to distinct disease susceptibilities and phenotypes that remain largely underexplored in African populations [5].

Induced pluripotent stem cells (iPSCs) can model genetic contributions to disease and help us understand the influence of specific cell populations to pathologic processes. iPSCs derived from dermal fibroblasts or peripheral blood mononuclear cells retain the donor’s genetic background [6]; have exponential proliferative capacity, making them a renewable resource; and can be engineered using CRISPR technology. Well-established protocols allow these iPSCs to be differentiated into diverse brain cell types, making them ideal for studying genetic influences on neurodegenerative diseases at the cellular level [7–11]. Since the advent of the technology used to generate iPSCs from somatic cells nearly two decades ago [12, 13], several global initiatives have emerged to support iPSC-based research in Europe [14], North America [15], and Asia [15–17]. Despite these advances, many iPSC collections still have significant gaps when it comes to the representation of diverse genetic ancestries [16]. For example, iPSCs from African populations are rare [18–20]. This underrepresentation severely limits the understanding of diseases and genetic variations pertinent to African populations [21].

To address this gap, we are building an open-access African somatic and stem cell bank as part of our African iPSC Initiative. Here, we describe the initial phase of this effort: 10 fibroblast and iPSC lines representing 5 Nigerian ethnic groups and both sexes. These lines were rigorously evaluated for pluripotency, genetic stability, and differentiation potential. Genomic profiling was performed in the iPSC for neurodegenerative risk genes, such as *APOE*, and polygenic risk scores (PRS) were calculated to estimate the overall burden of genetic factors associated with neurodegeneration in multiple ancestries and specific diseases. Using CRISPR/Cas9 genome editing, we further engineered frontotemporal dementia (FTD)-causing *MAPT* mutations (P301L and R406W). These iPSC lines, along with the fibroblasts used to generate them, represent the first dedicated open-access African Somatic and Stem Cell Bank to facilitate breakthroughs not only in the neurodegenerative field, but also in drug discovery and regenerative medicine.

## 2. Methods

### 2.1 Donor Eligibility and Consent

Ethical approval for this work was obtained from the Yobe State University Teaching Hospital Ethics Committee (Approval Reference: YSUTH/MAC/EA/077/VOL.III.297). Healthy adult donors were recruited based on eligibility criteria that required the absence of underlying chronic illnesses and neurological disorders, as well as negative screening results for common transmissible pathogens, including HIV and Hepatitis, to ensure the suitability of the samples for downstream iPSC applications. Before participation, all donors received detailed information about the study, including its purpose, procedures, potential risks, and benefits, both verbally and in writing. Written informed consent was obtained from all participants prior to sample collection. Consent included permission to derive, bank, and distribute iPSC lines for use in future research studies globally. Participants were assured that their data and samples would remain confidential and that their participation was voluntary.

### 2.2 Skin Biopsy Collection and Donor Demographics

A qualified surgeon at Yobe State University Teaching Hospital performed the procedure using a small surgical incision. Before the procedure, the forearm area was sterilized, and a local anaesthetic (lidocaine) was administered to numb the site. Following excision, the tissue samples were immediately rinsed twice in sterile phosphate-buffered saline (PBS) using two separate sterile 50 mL Falcon tubes to remove any contaminants. The biopsies were then transferred into sterile tubes containing Dulbecco’s Modified Eagle Medium (DMEM) supplemented with 1% (v/v) L-glutamine (L-Glu), 1% (v/v) penicillin/streptomycin (Pen/Strep), 2.5% (v/v) HEPES solution, and 10% (v/v) fetal calf serum (FCS). The biopsies were subsequently used to establish primary fibroblast cultures.

The cohort comprised ten donors representing five ethnic groups from Northern Nigeria: Kanuri, Hausa, Babur/Bura, Fulani, and Kare-Kare. Equal numbers of male and female participants were recruited, ranging from 18 to 60 years (**Table 1**).

**Table 1.** Human Fibroblasts.

| **Fibroblast Line**<br>**Name** | **Sex** | **Age at**<br>**Biopsy** | **Ancestry** | **APOE** |
| --- | --- | --- | --- | --- |
| BA-001 | M | 35-40 | Babur | 33 |
| BA-002 | M | 18-25 | Kanuri | 23 |
| BA-003 | M | 45-50 | Hausa | 23 |
| BA-004 | M | 18-25 | Kare Kare | 23 |
| BA-005 | M | 45-50 | Fulani | 33 |
| BA-006 | F | 60-65 | Kanuri | 33 |
| BA-007 | F | 25-30 | Babur | 33 |
| BA-008 | F | 18-25 | Hausa | 33 |
| BA-009 | F | 18-25 | Fulani | 23 |
| BA-010 | F | 18-25 | Kare Kare | 24 |

### 2.3 iPSC Generation

Human fibroblasts (**Table 1**) were transduced with non-integrating Sendai virus carrying OCT3/4, SOX2, KLF4, and cMYC (Life Technologies, A16518) in feeder- and serum-free conditions using mTeSR1 (STEMCell Technologies, 85850) as previously described [22]. Cells that showed morphological evidence of reprogramming were selected by manual dissection and maintained using mTeSR1. Three stable clones were expanded and banked for each donor line.

### 2.4 iPSC Characterization

Human iPSC lines (**Table 2**) were characterized using standard methods [22]. Authentication of the iPSC lines to their respective parental fibroblast was completed using short tandem repeat (STR) genotyping (**Supplemental Table 1**). Two clones for each donor line were analyzed for pluripotency markers (SOX2, SSEA4, OCT4, TRA-1-60) by immunocytochemistry and qPCR and for chromosomal abnormalities by G-band karyotyping (**Figure 2**; **Supplemental Figures 1-10**). Cell lines were confirmed to be free of mycoplasma. Detailed methods for each modality are described below.

**Table 2.**
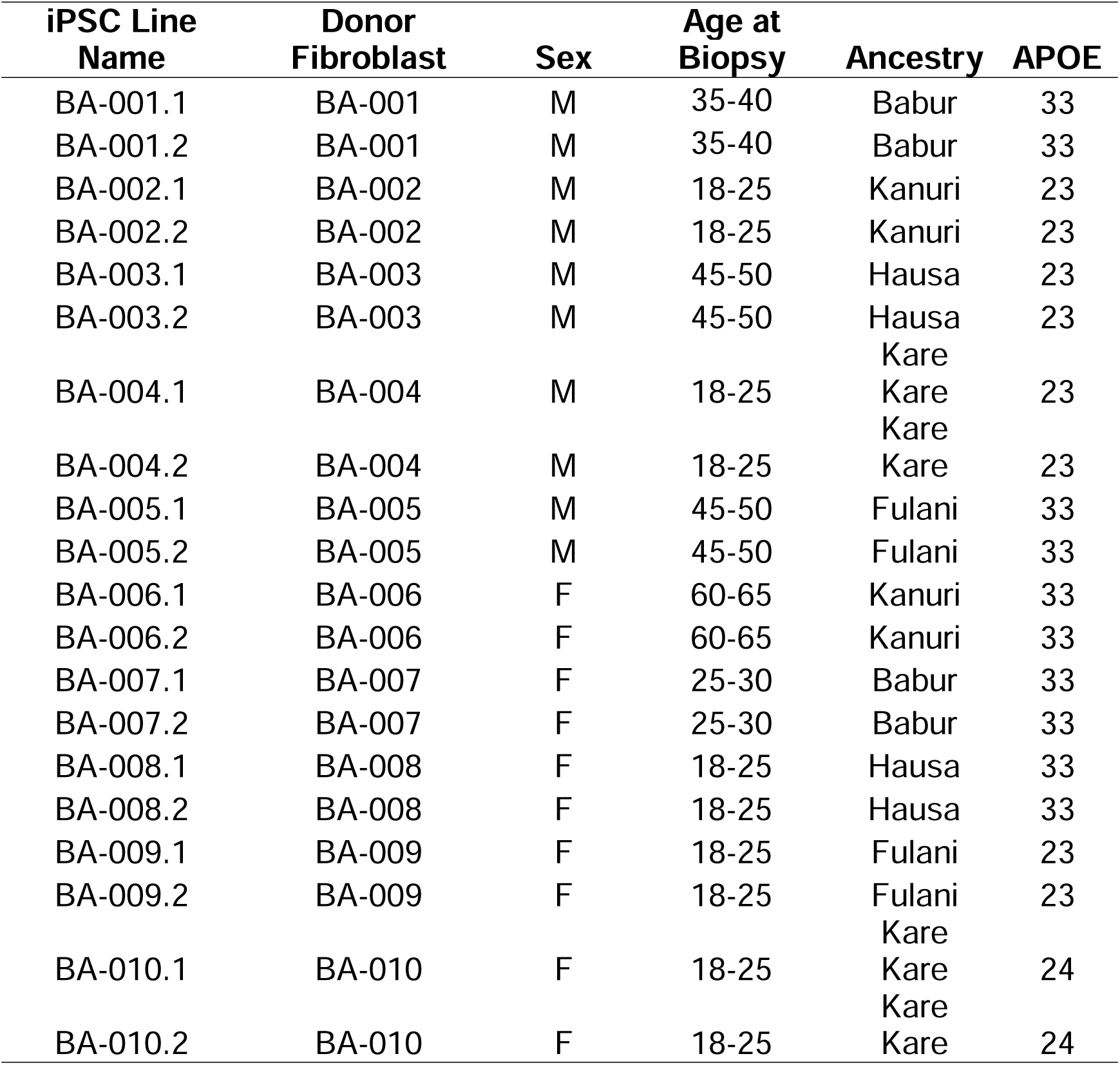
Human iPSC.

### 2.5 APOE Genotyping

Genomic DNA (gDNA) was extracted from cell pellets using a cell lysis solution (0.05% SDS, 0.1M Tris-Buffer, 0.1M EDTA in Milli-Q Water) as well as a protein precipitation solution (8M Ammonium Acetate). Cell lysis suspension was spun down for 1 minute at 16000*xg*. Supernatant was transferred to a solution of 100% isopropanol to isolate gDNA. gDNA pellets were washed with 70% ethanol and dissolved in 1X Tris-EDTA (TE) Buffer (G Biosciences, R025). *APOE* genotyping was completed by qPCR using an ABI Real Time Taqman SNP Genotyping assay. TaqMan probes used for allelic determination included rs429358 (C_3084793_20) and rs7412 (C_904973_10). The genotypes are defined by combining the results for both rs429358 and rs7412.

### 2.6 iPSC Culture

Human iPSCs were cultured in mTeSR1 on Cultrex Basement Membrane Extract [BME] -coated tissue culture-treated plates (R&D Systems, 3532-010-02). For routine passaging and unless otherwise noted below, iPSCs were dissociated with Accutase (STEMCell Technologies, 07922) for 3 minutes. Dissociated cells were collected in PBS and centrifuged at 750 rpm for 3 minutes. After medium was aspirated, the cells were plated on new BME-coated plates in mTeSR1. iPSCs were maintained with less than 5% spontaneous differentiation and were cryopreserved in mTeSR1 supplemented with 10% dimethyl sulfoxide and 40% FBS. iPSCs are karyotyped every 20 passages to ensure clones maintain stable genomes. All cell lines were confirmed to be mycoplasma-free using the MycoAlert mycoplasma detection kit (Lonza, LT07-710) according to the manufacturer’s instructions.

### 2.7 iPSC Genomic Editing

Human iPSCs underwent genomic editing via ribonucleoprotein (RNP). Three million cells were prepared by applying Accutase for ten minutes to achieve dissociation into single cells. Cells were then exposed to SNP specific crRNA and tracrRNA at a final concentration of 100µM, RNP Cas9 at a final concentration of 6.4µg/µL, and a donor oligonucleotide at a final concentration of 100µM. Cells then underwent electroporation from Lonza 4D-Nucleofector X unit and were plated in DMEM supplemented with 10% FBS. Five days post-nucleofection, cells were serial diluted and picked upon reaching 50-100 µm in size. Cells were then screened via Next Generation Sequencing (NGS) for introduction of the SNP of interest (*MAPT* P301L or *MAPT* R406W).

### 2.8 Immunocytochemistry

Cells were washed and fixed with 4% paraformaldehyde (Sigma-Aldrich, St. Louis, MO, USA). Primary and secondary antibodies were diluted in 3% bovine serum albumin. The following antibodies were used: SOX2 (1:500; Cell Signaling Technologies, 3579), SSEA4 (1:500; Cell Signaling Technologies, 4755), TRA-1-60 (1:1000; Cell Signaling Technologies, 4746), OCT4 (1:500; Cell Signaling Technologies, 2840), Donkey Anti-Rabbit Alexa Fluor 594 (1:250; Life Technologies, A-21207), Goat Anti-Mouse Alexa Fluor 488 (1:250; Life Technologies, A-10680), Donkey Anti-Rabbit Alexa Fluor 488 (1:250; Life Technologies, A-21206), and Goat Anti-Mouse Alexa Fluor 594 (1:250; Life Technologies, A-11005). Nuclei were counterstained with 4′,6-diamidino-2-phenylindole (DAPI; Life Technologies). Images were acquired on a Nikon Eclipse 80i fluorescence microscope (Nikon Instruments, Melville, NY, USA) using LMS software.

### 2.9 qPCR

RNA was extracted from cell pellets with a RNeasy kit (QIAGEN, 74106), following the manufacturer’s protocol. Extracted RNA (10 μg) was converted to complementary DNA (cDNA) by PCR using the High-Capacity cDNA Reverse Transcriptase Kit (Life Technologies, 4368813). Gene expression was measured in iPSCs for *SOX2*, *POU5F1*, *LIN28A*, *NANOG*, *B3GALT5*, and *PODXL* using qPCR as previously described [23]. Primers specific to Sendai virus (*SEV*) were included to evaluate the presence of virus remaining in the isolated clones. Primers specific to *GAPDH* were used as a control.

### 2.10 Karyotyping

Chromosomal abnormalities were assessed by G-band karyotyping.

### 2.11 Trilineage Differentiation

The capacity to form cell types from the three germ layers was confirmed by differentiating cells using STEMdiff Trilineage Differentiation Kit (STEMCell Technologies, 05230). Briefly, cells were plated according to manufacturer’s protocol and supplemented with either STEMdiff Trilineage Endoderm Medium, STEMdiff Trilineage Ectoderm Medium, or STEMdiff Trilineage Mesoderm Medium. Cells were harvested after five days, and qPCR was performed as previously described to assess expression of germ layer-specific markers (Ectodermal Markers: *TUBB3* and *Pax6*; Endoderm Markers: *FOXA2* and *CXCR4*; Mesodermal Markers: *Desmin* and *SMA*). Primers specific to *GAPDH* and *Fibronectin* were used as endogenous controls and fibroblast markers, respectively.

### 2.12 Genomic Analyses

gDNA was isolated as described above and resuspended at a dilution of at least 50 ngl/ul were analyzed by an Infinium Global Screening Array-24 v.30 GWAS Chip. Quality control (QC) of the GWAS data was performed using PLINK 1.9 [24] and plinkQC 0.3.4 [25]. Low-quality single nucleotide polymorphisms (SNPs) were filtered using the following criteria: (1) SNP missing rate > 1%; (2) minor allele frequency (MAF) <5%; and (3) Hardy–Weinberg P value <1 × 10^−6^ (**Supplemental Table 2**). Samples were verified for sex inconsistencies by matching phenotypic data with genetic sex imputation from the X-chromosomal heterozygosity rate. Identity-by-descent (IDB) analysis was performed to identify pairs of fibroblasts and iPSC lines from the same donor and to estimate the genetic relationships between individuals. SNPs from *APOE* genotyping (rs7412 and rs429358) were added manually to the GWAS results.

After QC, bcftools 1.21 [26] was used to convert SNP files to VCF files, following the data phasing with Eagle v2.4 [27] and the imputation of un-genotyped variants using Michigan Imputation Server 2 (version 2.0.6) [28] with 1000 Genomes Phase 3 v5 as a reference panel [29]. Imputed SNPs with R^2^ < 0.3 were removed and used to estimate the global ancestry of samples by running PCA on 1000 Genomes Phase 3 v5 reference data and projecting the calculated principal components to the target dataset using default parameters. The FRAPOSA tool [30] was used to estimate the distance to the population in the reference panel.

### 2.13 Polygenic Risk Scores (PRS)

PRS for Alzheimer’s disease (AD) and Parkinson’s disease (PD) were calculated with pgsc_calc pipeline [31–34]. Score files with GWAS risk estimates were downloaded from the PGS catalog [31]. GWAS summary statistics, PGS003956 [35] and PGS000903 [36], were used as a reference for AD and PD polygenic score (PGS) calculations, respectively. A comparison dataset of African population genotypes was obtained from 1000 Genomes Phase 3 v5 data by selecting donors with African global ancestry. PRS excluding *APOE* regions were calculated separately by excluding SNPs within the 200Kb region surrounding the *APOE* gene locus (hg19 positions – chr19:45,211,000-45,613,000).

### 2.14 RNA Sequencing

iPSCs were resuspended in 200 µL of 50:1 homogenization solution: 1-Thioglycerol solution. After addition of 200 µL of Promega lysis buffer, the samples were transferred to the appropriate well of the Maxwell RSC cartridge. DNase solution was added to each cartridge. TapeStation 4200 System (Agilent Technologies) was used to perform quality control of the RNA concentration, purity, and degradation based on the estimated RNA integrity Number (RIN) and DV200. Samples were sequenced by an Illumina HiSeq 4000 Systems Technology with a read length of 1×150 bp and an average library size of 36.5 ± 12.2 million reads per sample.

Identity-by-Descent (IDB) [37] and FastQC [38] analyses were performed to confirm sample identity. STAR (v.2.6.0) [39] was used to align the RNA sequences to the human reference genome: GRCh38.p13 (hg38). The quality of RNA alignment was evaluated using sequencing metrics such as read distribution, ribosomal content, and alignment quality in Picard (v.2.8.2).

Salmon (v. 0.11.3) [40] was used to quantify the expression of the genes annotated within the human reference genome used in this project (GRCh38.p13). Protein coding genes were selected for downstream analyses. Principal component analysis (PCA) was calculated based on the 500 most variably expressed genes obtained from 19,957 protein-coding genes using regularized logarithm transformation (rlog) counts. PCA and histogram plots were created using the ggplot2 package (V.3.3.6) [41].

## 3. Results

### 3.1 Community Engagement and Development of African Cell Bank

Africa’s unparalleled genetic diversity is vastly underrepresented in current cellular models [21]. We set out to address this critical gap by building an open-access African somatic and stem cell bank. Cells in this bank represent indigenous Africans from Nigeria, where we have leveraged decades-long connections and collaboration with communities to facilitate engagement with this initiative [42–44].

To achieve broad community participation, we conducted extensive outreach activities, including radio campaigns, across northeastern Nigeria to raise awareness of the study and recruit participants. Informed consent was secured from healthy donors who met eligibility criteria (see Methods). Skin biopsies were performed on 10 donors (5 males and 5 females) representing five ethnic groups in the region: Kanuri, Hausa, Babur/Bura, Fulani, and Kare-Kare (**Figure 1; Table 1**). Each sample was assigned a unique identifier to maintain anonymity. The ethnic donors recruited in the Yobe State are broadly represented across northern Nigeria and, in some cases, extend beyond Nigeria’s borders (**Figure 1A**). The Hausa and Fulani populations are widespread across West Africa, while the Kanuri population has historical ties to the Kanem-Bornu Empire, with communities spanning regions of Cameroon, Niger, and Chad.

**Figure 1:**
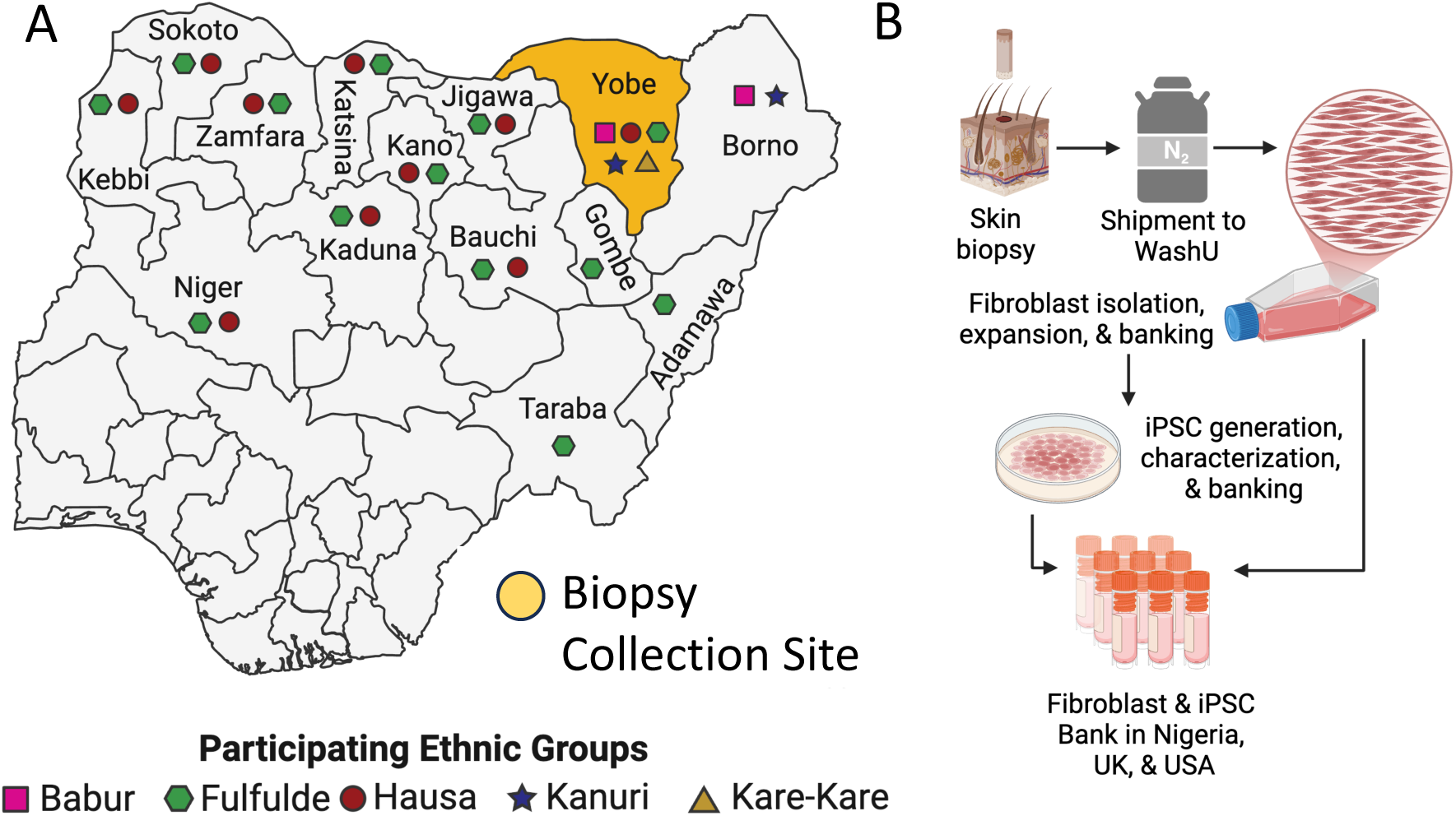
Establishing an African Somatic and Stem Cell Bank. A. Map of Nigeria Indicating biopsy collection site (yellow) in the state of Yobe. Ethnic groups participating in the study indicated along with their regional distribution. Pink square, Babur. Green hexagon, Fulfude. Red circle, Hausa. Blue star, Kanuri. Yellow triangle, Kare-Kare. B. Workflow to transform skin biopsies into fibroblasts and induced pluripotent stem cells (iPSC).

Skin biopsies were processed to obtain primary fibroblast cultures and banked to ensure accessibility for future research. Continued collection of fibroblast and erythroid progenitor cells from healthy and AD donors will continue to expand the somatic cell bank in the future. Together, these efforts represent a pivotal step in the African iPSC Initiative: fostering community participation and trust while establishing a somatic cell resource that will support future scientific discoveries and global research collaborations.

### 3.2 Generation of iPSCs for an African Stem Cell Bank

Fibroblast cultures from the 10 donor lines were reprogrammed into iPSCs using non-integrating Sendai virus vectors carrying OCT3/4, SOX2, KLF4, and cMYC (**Figure 1B; Table 2**). Phase-contrast microscopy revealed that colonies from all the iPSCs have a high nuclear-to-cytoplasmic ratio with prominent nucleoli, grow in colonies, and have well-defined borders, characteristic of iPSCs (**Figure 2A**). RNA expression of key pluripotency markers, including *SOX2*, *POU5F1*, *LIN28A*, *NANOG*, and *PODXL*, were increased (**Figure 2B**, **Supplemental Figures 1-10**). Expression levels of pluripotency markers varied slightly across iPSC donor lines (**Supplemental Figures 1-10**), possibly due to differences in genetic background across donors, which has been reported to be the major driver of phenotypic variability in iPSCs [45]. Stability of iPSC clones is dependent upon expression of endogenous pluripotent genes and elimination of residual exogenous Sendai virus. Primers specific to Sendai virus illustrated clearance of residual virus in the reprogrammed clones (**Figure 2B; Supplemental Figures 1– 10**). Immunostaining with antibodies for the pluripotency markers SOX2, OCT4, SSEA4, and TRA-1-60, further confirmed the pluripotent identity of the iPSC lines (**Figure 2C; Supplemental Figures 1–10**).

**Figure 2:**
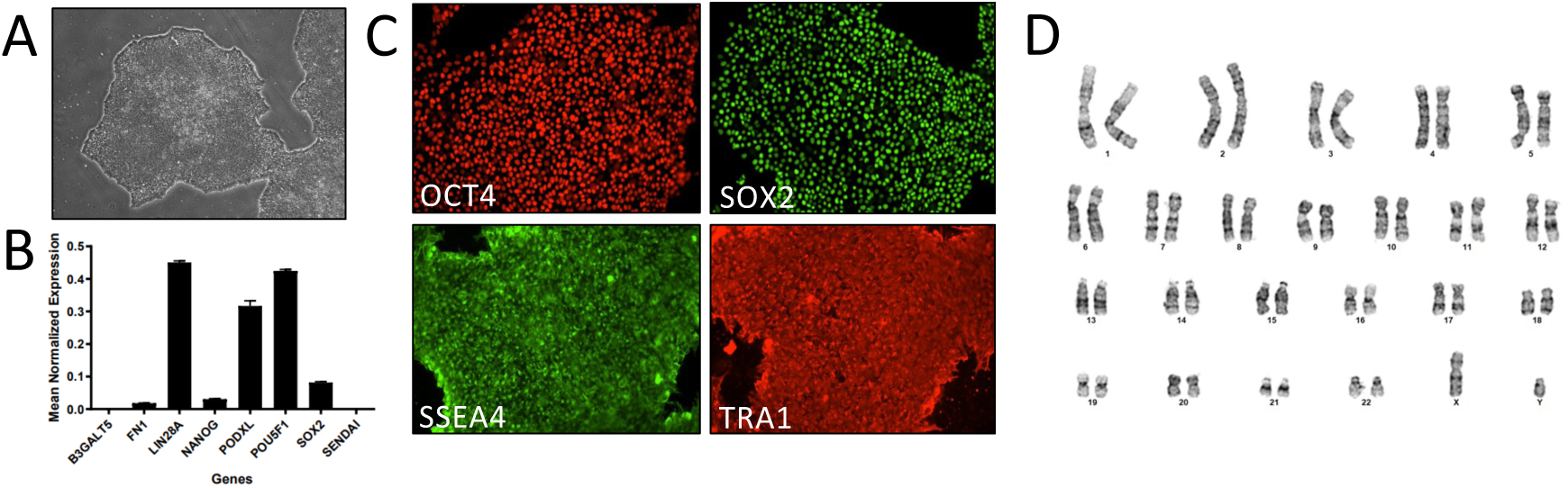
Characterization of iPSC lines from African Somatic and Stem Cell Bank. Representative characterization data featuring iPSC BA-001.1 A. Phase-contrast images showing cultured iPSCs. B. qPCR for pluripotency markers. C. Immunocytochemistry for pluripotency markers OCT4, SOX2, SSEA4, and TRA-1-60. D. G-band karyotyping reveals no chromosomal abnormalities. See Supplemental Figures 1-10 for characterization of all iPSC described in this study.

To evaluate the genomic integrity of the iPSC lines, karyotyping analysis was conducted, revealing no detectable chromosomal abnormalities (**Figure 2D; Supplemental Figures 1–10**). STR profiling illustrates that all the iPSC clones are a genetic match for their parental fibroblast lines (**Supplemental Table 1**). Together, these analyses demonstrate that the iPSC lines are stable and robust for downstream applications.

### 3.3 Genomic and Molecular Characterization of iPSCs for an African Stem Cell Bank

To maximize utility of the cell bank, genomic analyses were performed to evaluate common variants (**Figure 3A**). After QC (see Methods), genotype information from 333,080 SNPs was used to evaluate genetic ancestry of the iPSC lines. PCA was performed by comparing each iPSC line with the 1000 Genomes reference ancestry panels. PCA revealed that the iPSC lines (gray circles) cluster closest to the reference data from African/African American ancestry (**Figure 3B**). Thus, the genomic background of the iPSC is enriched for African/African American ancestry.

**Figure 3:**
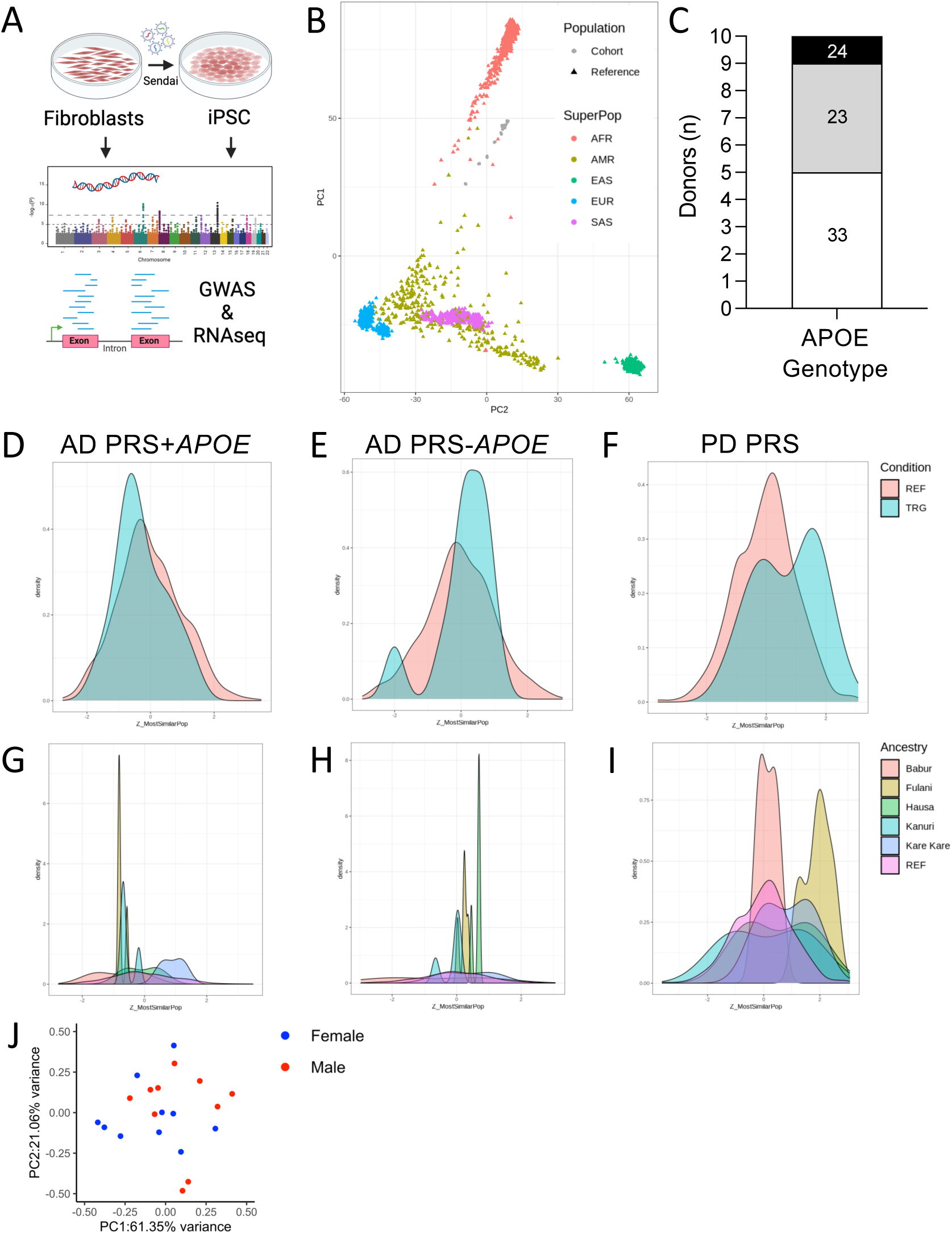
Genomic Profiling of iPSC lines from African Somatic and Stem Cell Bank. A. Workflow. B-I. Genetic analyses of iPSC. B. Principal component analysis (PCA) of common genetic variants measured in unique iPSC lines (gray, cohort) compared to 1000 Genomes reference: Africans (AFR), Admixed Americans (AMR), East Asians (EAS), Europeans (EUR), South Asians (SAS). C. Frequency of *APOE* genotypes among iPSC lines. D-I. Polygenic Risk Scores (PRS). D-E. Distribution of PRS represented using reference (salmon, REF) and iPSC samples (cyan, TRG). G-H. Distribution of PRS represented based on Nigerian ethnic group. D, G. PRS for Alzheimer’s disease (AD) including the *APOE* gene locus. E, H. PRS for AD excluding the *APOE* gene locus. F, I. PRS for Parkinson’s disease (PD). J. Principal component analysis of transcriptomic data generated from iPSCs (500 most variable gene transcripts). Blue, female. Red, male.

The three major *APOE* isoforms*, APOE2, APOE3,* and *APOE4*, contribute to AD risk by varying degrees*. APOE4* has been identified as a major risk factor for AD, while *APOE2* is considered protective [46]. Yet, the penetrance of *APOE* genotypes differs across ancestral backgrounds [47]. *APOE33* was the most common genotype among the cell lines (n=5, **Figure 3C**). *APOE*33 is considered neutral across ancestral backgrounds [46]. *APOE*23 (n=4) is associated with reduced AD risk (Odds Ratio (OR) 0.6), and *APOE*24 (n=1) is associated with increased AD risk (OR 2.6) (**Figure 3C**)[48]. Thus, the iPSC collection represents *APOE* genotypes that capture the spectrum of AD risk.

While *APOE* is a major risk factor for AD, additional common variants contribute to disease risk [46]. The majority of these variants have small effects on disease risk, and thus, genetic risk profiles are optimally interpreted by combining multiple risk variants to generate a score (PRS) [49–51]. PRSs are informative for a number of neurodegenerative diseases where complex genetic architecture underlies risk for sporadic disease. Leveraging genotype data from the 333,080 SNPs, we generated PRS for each of the lines using the polygenic score catalog (**Figure 3D-I, Supplemental Table 3**) [31]. Distribution of PRS was plotted relative to a reference dataset (donors with African global ancestry in 1000 Genomes Phase 3 v5; **Figure 3D-I**). PRSs constructed from the summary statistics of five AD GWAS[35] were plotted for the iPSC lines, including the *APOE* locus (**Figure 3D, Supplemental Table 3**) and excluding the *APOE* locus (**Figure 3E, Supplemental Table 3**). PRSs were normally distributed, suggesting this collection captures the spectrum of AD PRSs. Additionally, PRSs were calculated using the summary statistics from 17 GWAS for PD[36] and plotted for the iPSC lines (**Figure 3F**). PD PRSs were bimodally distributed among the iPSC (**Figure 3F, Supplemental Table 3**). As genetic ancestry contributes to common variants that confer disease risk and, in turn, PRS, we sought to evaluate the distribution of PRSs for AD and PD based on the ethnic group of the iPSC (**Figure 3G-I**). Frequency of high and low PRSs differ based on the ethnic group; however, because the sample numbers remain small, we cannot make biological conclusions about these observations.

Molecular profiling of the iPSC lines using bulk RNA sequencing was also performed. By comparing the 500 most variably expressed genes, we find that the iPSCs are distributed in a pattern consistent with prior observations that donor background is the largest contributor to transcriptomic variability[45]. Additionally, transcriptomic profiles do not differ based on sex (**Figure 3J**). Together, the genomic and molecular profiles provide a strong foundation for selection and use of these lines for future studies.

### 3.4 Differentiation Potential of the iPSCs for an African Stem Cell Bank

A key component of a high-quality iPSC line is its capacity to differentiate into cells within the three major germ layers. To evaluate the differentiation capacity of the newly generated iPSC lines, we performed trilineage differentiation assays using the STEMdiff Trilineage Differentiation Kit. Expression analysis revealed robust expression of markers in each of the three lineages. Expression of *PAX6* and *TUBB3*, markers associated with neuroectodermal lineages, confirm ectodermal differentiation (**Supplemental Figures 1–10**). Mesodermal differentiation was validated by the strong expression of *SMA* and *DES*, albeit to a lesser extent (**Supplemental Figures 1–10**). Endodermal differentiation was confirmed with expression of *FOX2A* and *CXCR4* (**Supplemental Figures 1–10**). All 20 iPSC clones (from 10 donor lines) successfully differentiated into cells in each of the three germ layers, though expression levels for lineage markers differed slightly across donors. This is consistent with prior reports of genetic background-driven differences in differentiation efficiency [45]. Together, we demonstrate that all the iPSC lines can differentiate into derivatives of the three germ layers, highlighting their utility for broad applications.

### 3.5 Genome Engineering to Model Genetic Causes of FTD in Diverse Backgrounds

To begin to expand the utility of these iPSC lines for researchers in the dementia field, we employed CRISPR/Cas9-mediated genome editing to introduce pathogenic *MAPT* mutations into the newly characterized iPSC lines (**Figure 4A; Table 3**). This effort aligns with prior endeavours to establish genetically engineered iPSC models for studying tau-related neurodegeneration [8, 11]. The iPSC line BA-001.1 was selected to introduce one of two well-characterized *MAPT* mutations—P301L or R406W—both of which are associated with frontotemporal lobar degeneration and other tauopathies [52–55]. The *MAPT* P301L mutation is one of the most common variants worldwide associated with FTD, leading to behavioral disturbances, aphasia, cognitive impairment, and parkinsonism [56]. Similarly, the *MAPT* R406W mutation has been identified in multiple families worldwide. Interestingly, *MAPT* R406W carriers exhibit a clinical phenotype that mimics AD, including progressive cognitive impairment [57–59]. CRISPR/Cas9 editing was performed with reagents previously described for engineering *MAPT* mutations on a wild-type background [8].

**Figure 4:**
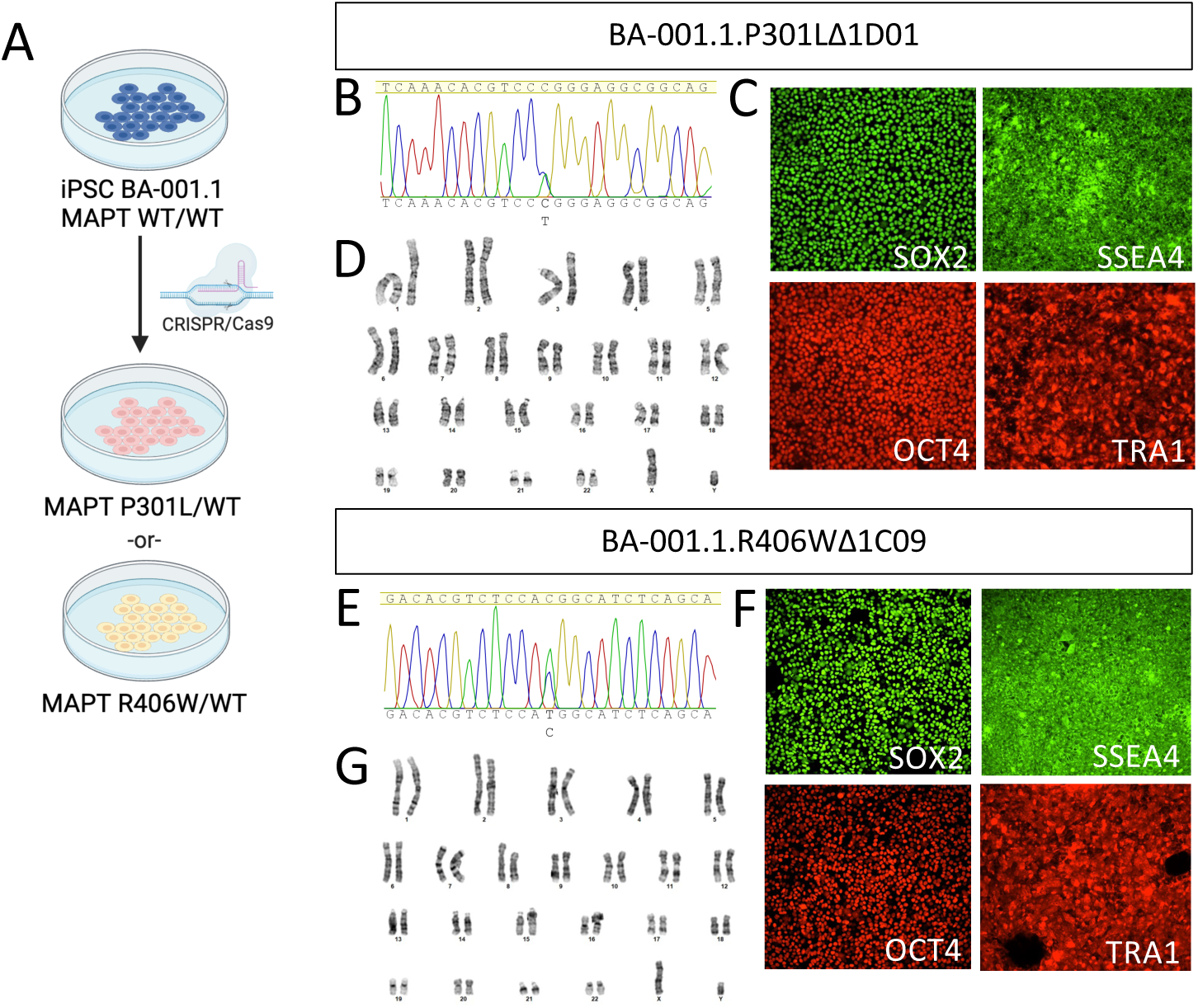
Genome engineering of the MAPT locus in iPSC BA-001.1. A. Workflow. B-D. BA-001.1 was edited from *MAPT* WT/WT to *MAPT* P301L/WT using CRISPR/Cas9. Characterization for one of the resulting clones: BA-001.1.P301L Δ1D01. B. Sanger sequencing illustrating the heterozygous engineered SNP. C. Immunocytochemistry for pluripotency markers OCT4, SOX2, SSEA4, and TRA-1-60. D. G-band karyotyping reveals no chromosomal abnormalities. E-F. BA-001.1 was edited from *MAPT* WT/WT to *MAPT* R406W/WT using CRISPR/Cas9. Characterization for one of the resulting clones: BA-001.1.R406W Δ1C09. E. Sanger sequencing illustrating the heterozygous engineered SNP. F. Immunocytochemistry for pluripotency markers OCT4, SOX2, SSEA4, and TRA1. G. G-band karyotyping reveals no chromosomal abnormalities.

**Table 3.** Genome editing of MAPT locus to model Frontotemporal Dementia.

| **iPSC** | **iPSC Donor Line** | **Donor Fibroblast** | **Engineered Genotype** | **Sex** |
| --- | --- | --- | --- | --- |
| BA-001.1.P301LΔ1D01 | BA-001.1 | BA-001 | MAPT P301L/WT | M |
| BA-001.1.R406WΔ1C09 | BA-001.1 | BA-001 | MAPT R406W/WT | M |

Two clones were fully characterized for each *MAPT* mutation (**Figure 4** and **Supplemental Figures 11-12**). Sanger sequencing chromatograms confirmed heterozygous single-nucleotide substitutions at the expected *MAPT* loci for both the P301L (**Figure 4B**; BA-001.1.P301LΔ1D01) and R406W mutations (**Figure 4E**; BA-001.1.R406WΔ1C09). We next evaluated the pluripotency and genomic stability of the edited lines using immunocytochemistry and karyotyping. iPSC BA-001.1.P301LΔ1D01 and iPSC BA-001.1.R406WΔ1C09 illustrated robust expression of key pluripotency markers, SOX2, SSEA4, OCT4, and TRA-1-60 (**Figure 4C** and **4F**), consistent with the unedited iPSCs. Additionally, G-band karyotyping analysis revealed no detectable chromosomal abnormalities in the edited clones, demonstrating that genomic integrity was preserved following the editing process (**Figure 4D** and **4G**). Together, this mutant *MAPT* engineered series will be a valuable tool for elucidating the molecular mechanisms driving neurodegenerative tauopathies within the unique genetic context of African ancestry.

### 3.6. Accessing Data and Cells from the African Somatic and Stem Cell Bank

To facilitate global access to the fibroblast and stem cell resources, we have implemented a decentralized biobanking system for cell distribution. The cells described here are stored at the Biomedical Science Research Training Centre (BioRTC) at Yobe State University, Damaturu, Nigeria, which serves as the primary repository (www.BioRTC.com). To promote global distribution, cells are also stored at the Knight Alzheimer Disease Research Center (Knight ADRC) Biomarker Core at Washington University School of Medicine, USA, and at the Cell Bank at the School of Life Sciences, University of Sussex, UK. Requests for cell lines are managed centrally via the primary repository at BioRTC. Approved requests will require a fully executed universal Material Transfer Agreement (MTA) and will be fulfilled by the repository geographically nearest to the requester to minimize shipping times. Our initiative will support diverse research fields, foster global collaboration, and accelerate innovation in understanding and treating human diseases.

## 4. Discussion

Here, we described the generation of 10 deeply phenotyped iPSC lines from five ethnic groups of Northern Nigeria as the initial phase of the African iPSC Initiative to derive and distribute iPSC models from indigenous Africans to the international scientific community. Africans represent the most genetically diverse population globally, harboring a vast array of common variants considered rare in other populations [2, 60]. This genetic diversity presents a rich resource for studying disease mechanisms, drug discovery, and regenerative therapies [61, 62]. Yet, cellular models that capture this diversity are rare [16, 21]. The collection described here builds upon our recent report of the first iPSC line from indigenous Nigerians [19].

### 4.1 Open Access African Somatic and Stem Cell Bank

The availability of population-specific somatic cells and iPSCs is critical for elucidating the molecular underpinnings of diseases across diverse populations. Globally, major initiatives have emerged to generate and bank iPSCs. For example, in Europe, the European Bank for Induced Pluripotent Stem Cells (EBiSC) serves as a centralized repository for standardized iPSC lines [14]. In the United States, institutions such as the California Institute for Regenerative Medicine (CIRM), the Coriell Institute for Medical Research, the WiCell Research Institute, the National Centralized Repository for Alzheimer’s Disease and Related Dementias (NCRAD), and the NIH Center for Alzheimer’s and Related Dementias (CARD) iPSC Neurodegenerative Disease Initiative (iNDI) via The Jackson Laboratory play key roles in distributing well-characterized iPSC lines, particularly for neurodegenerative disease research [15, 63, 64]. In Asia, the RIKEN BioResource Research Center and Center for iPS Cell Research and Application (CiRA) in Japan, National Stem Cell Bank of Korea, and Human Disease iPSC Consortium Resource Center play key roles in distributing iPSC lines [15–17]. However, among the iPSC lines with assigned ethnicities in these repositories, only a small percentage represent Black or African ancestry, primarily derived from African American donors [21]. Populations of African descent living outside the continent do not fully capture the immense genetic diversity of the ethnolinguistic groups across Africa [21, 61, 62, 65].

To address this shortcoming, we have established the first African Somatic and Stem Cell Bank, which contains a collection of somatic cells and iPSC lines. Each cell line in this resource has undergone rigorous quality control, including pluripotency testing, karyotyping, and STR profiling to confirm iPSC identity, genetic integrity, and stability. These iPSCs exhibit robust differentiation potential into derivatives of the three germ layers, demonstrating their versatility for diverse applications. These iPSCs can support a broad range of research applications, including disease modelling, drug discovery, and regenerative medicine, thus enabling breakthroughs that better reflect the genetic backgrounds of African populations.

The fibroblasts in our biobank offer unique advantages for translational research. Fibroblasts can be directly reprogrammed into induced neurons without transitioning through an iPSC state [66–71]. This approach not only reduces the time required for cell reprogramming but also retains the epigenetic state of the biopsied cells, which are often reset during iPSC generation [72]. Retaining these epigenetic features is particularly important for studying age-related diseases, as it allows these induced neurons to reflect the aging phenotype of the donor [71]. Recent work revealed that induced neurons express tau isoforms at both the transcript and protein levels, closely mirroring what is observed in the adult human brain [68]. This makes such neurons valuable for investigating the onset and progression of neurodegenerative diseases [68, 73]. Thus, our fibroblast resource offers a powerful platform for generating induced neurons to explore critical aspects of neurodegeneration within the context of African genetic background. Overall, our cell biobank sets a critical precedent for future biobanking efforts in Africa and serves as a critical resource for diverse fields requiring access to cells from African genetic backgrounds.

### 4.2 Genome Editing to Study Tauopathies

In line with global efforts to develop disease-specific iPSC models and our longstanding interest in tauopathies, we used CRISPR/Cas9-mediated genome editing to generate iPSC lines carrying well-characterized *MAPT* mutations, P301L or R406W, which are linked to FTD [52–55]. These mutant lines retain key pluripotency markers and exhibit normal karyotypes, demonstrating their suitability for investigating tau-related pathologies. While their intrinsic value for understanding disease mechanisms is significant, their utility is further enhanced when compared with isogenic lines from other populations [8, 11, 74–77], which can enable a comprehensive examination of ancestry-related factors influencing the cellular mechanisms underlying tauopathies. Moreover, given the population-level differences in *MAPT* haplotypes, this resource offers a unique opportunity to examine whether ancestry-specific genetic variations modify tau expression, splicing, or aggregation in tauopathies. Thus, by integrating these edited lines into our biobank, we further extend the utility of this resource for researchers studying tauopathies and to support drug discovery pathways.

### 4.3 Global Collaboration and Distribution of iPSC Lines

To address the absence of a local facility for generating and banking iPSCs in Nigeria, the Biomedical Science Research Training Centre (BioRTC; www.BioRTC.com) was established at Yobe State University, Nigeria, in 2021 [43]. This center, supported by the Yobe State Government, now serves as the repository and distribution hub for the generated iPSC lines and their precursor somatic cells, with sub-repositories established at existing biobanks at Washington University School of Medicine and the University of Sussex. All ten iPSC lines, the *MAPT*-mutant isogenic pairs, and a previously generated iPSC line from an adult Nigerian donor [19], have been banked at this newly established African Somatic and Stem Cell Bank.

Given that broad consent for global distribution was obtained from donors, these iPSC lines and derivatives are available to the global scientific community under a Universal MTA. Our vision is that this resource—and additional lines currently in development—will provide a deeply characterized and genetically diverse collection of iPSCs derived from indigenous Africans. This will foster global collaboration, drive breakthroughs in the study of human diseases beyond neurodegenerative disorders, and accelerate drug discovery by enabling research in cells derived from African genetic backgrounds.

### 4.4 Concluding Remarks

The African iPSC Initiative marks a transformative step in addressing the global underrepresentation of African populations in biomedical research. The iPSCs and their derivatives generated through this effort offer unparalleled potential for advancing our understanding of human diseases, particularly tauopathies, and for accelerating drug discovery in a historically underserved population. Future efforts aim to expand this resource by including iPSCs from a broader spectrum of African ethnic groups and generating disease-specific lines, such as those for AD, to further support research on neurodegenerative and other diseases.

## Data Availability

All data produced in the present study are available upon reasonable request to the authors

https://www.biortc.com

## Acknowledgments

We would like to thank the research subjects and their families who generously participated in this study. We thank Torri Ball for careful review and thoughtful discussion of the study. We thank the Genome Engineering & Stem Cell Center (GESC@MGI) at Washington University in St. Louis School of Medicine for iPSC reprogramming and editing services. This work was supported by access to equipment made possible by the Hope Center for Neurological Disorders, the Neurogenomics and Informatics Center, and the Departments of Neurology and Psychiatry at Washington University School of Medicine. Diagrams were generated using BioRender.com.

## Consent Statement

Written informed consent was obtained from all participants prior to sample collection. Consent included permission to derive, bank, and distribute iPSC lines for use in future research studies globally.

## References

[1] Blommaert J. Linguistics diversity: Africa. 2007. p. 123–49.

[2] Campbell MC, Tishkoff SA. African genetic diversity: implications for human demographic history, modern human origins, and complex disease mapping. Annu Rev Genomics Hum Genet. 2008;9:403–33.

[3] Reitz C, Pericak-Vance MA, Foroud T, Mayeux R. A global view of the genetic basis of Alzheimer disease. Nature Reviews Neurology. 2023;19:261–77.

[4] Jonson C, Levine KS, Lake J, Hertslet L, Jones L, Patel D, et al. Assessing the lack of diversity in genetics research across neurodegenerative diseases: A systematic review of the GWAS Catalog and literature. Alzheimers Dement. 2024;20:5740–56.

[5] Weinberger DR, Dzirasa K, Crumpton-Young LL. Missing in Action: African Ancestry Brain Research. Neuron. 2020;107:407–11.

[6] Kyttälä A, Moraghebi R, Valensisi C, Kettunen J, Andrus C, Pasumarthy Kalyan K, et al. Genetic Variability Overrides the Impact of Parental Cell Type and Determines iPSC Differentiation Potential. Stem Cell Reports. 2016;6:200–12.

[7] Penney J, Ralvenius WT, Tsai L-H. Modeling Alzheimer’s disease with iPSC-derived brain cells. Molecular Psychiatry. 2020;25:148–67.

[8] Karch CM, Kao AW, Karydas A, Onanuga K, Martinez R, Argouarch A, et al. A Comprehensive Resource for Induced Pluripotent Stem Cells from Patients with Primary Tauopathies. Stem Cell Reports. 2019;13:939–55.

[9] Rivetti di Val Cervo P, Besusso D, Conforti P, Cattaneo E. hiPSCs for predictive modelling of neurodegenerative diseases: dreaming the possible. Nature Reviews Neurology. 2021;17:381–92.

[10] Wray S. Modelling neurodegenerative disease using brain organoids. Seminars in Cell & Developmental Biology. 2021;111:60–6.

[11] Karch CM, Hernández D, Wang JC, Marsh J, Hewitt AW, Hsu S, et al. Human fibroblast and stem cell resource from the Dominantly Inherited Alzheimer Network. Alzheimers Res Ther. 2018;10:69.

[12] Takahashi K, Tanabe K, Ohnuki M, Narita M, Ichisaka T, Tomoda K, et al. Induction of Pluripotent Stem Cells from Adult Human Fibroblasts by Defined Factors. Cell. 2007;131:861–72.

[13] Takahashi K, Yamanaka S. Induction of Pluripotent Stem Cells from Mouse Embryonic and Adult Fibroblast Cultures by Defined Factors. Cell. 2006;126:663–76.

[14] De Sousa PA, Steeg R, Wachter E, Bruce K, King J, Hoeve M, et al. Rapid establishment of the European Bank for induced Pluripotent Stem Cells (EBiSC) - the Hot Start experience. Stem Cell Research. 2017;20:105–14.

[15] Gurwitz D, Steeg R. Enriching iPSC research diversity: Harnessing human biobank collections for improved ethnic representation. Drug Development Research. 2024;85:e22227.

[16] Ghosh S, Nehme R, Barrett LE. Greater genetic diversity is needed in human pluripotent stem cell models. Nature Communications. 2022;13:7301.

[17] Huang C-Y, Liu C-L, Ting C-Y, Chiu Y-T, Cheng Y-C, Nicholson MW, et al. Human iPSC banking: barriers and opportunities. Journal of Biomedical Science. 2019;26:87.

[18] Mura M, Pisano F, Stefanello M, Ginevrino M, Boni M, Calabrò F, et al. Generation of two human induced pluripotent stem cell (hiPSC) lines from a long QT syndrome South African founder population. Stem Cell Research. 2019;39:101510.

[19] Muhammad Z, Brown PW, Babazau L, Alkhamis AI, Goni BW, Nggada HA, et al. Generation of an induced pluripotent stem cell line (BIORTCi001-A) from a healthy adult indigenous Nigerian participant. Stem Cell Research. 2024;80:103503.

[20] Naidoo J, Hurrell T, Scholefield J. The generation of human induced pluripotent stem cell lines from individuals of Black African ancestry in South Africa. Stem Cell Research. 2024;81:103534.

[21] Hurrell T, Naidoo J, Ntlhafu T, Scholefield J. An African perspective on genetically diverse human induced pluripotent stem cell lines. Nature Communications. 2024;15:8581.

[22] Feigin VL, Nichols E, Alam T, Bannick MS, Beghi E, Blake N, et al. Global, regional, and national burden of neurological disorders, 1990&#x2013;2016: a systematic analysis for the Global Burden of Disease Study 2016. The Lancet Neurology. 2019;18:459–80.

[23] Karch CM, Kao AW, Karydas A, Onanuga K, Martinez R, Argouarch A, et al. A Comprehensive Resource for Induced Pluripotent Stem Cells from Patients with Primary Tauopathies. Stem Cell Reports. 2019;13:939–55.

[24] Purcell S, Neale B, Todd-Brown K, Thomas L, Ferreira MA, Bender D, et al. PLINK: a tool set for whole-genome association and population-based linkage analyses. Am J Hum Genet. 2007;81:559–75.

[25] Meyer HV. meyer-lab-cshl/plinkQC: plinkQC version 0.3.4 (v0.3.4). Zenodo. 2021.

[26] Danecek P, Bonfield JK, Liddle J, Marshall J, Ohan V, Pollard MO, et al. Twelve years of SAMtools and BCFtools. Gigascience. 2021;10.

[27] Loh PR, Danecek P, Palamara PF, Fuchsberger C, Y AR, H KF, et al. Reference-based phasing using the Haplotype Reference Consortium panel. Nat Genet. 2016;48:1443–8.

[28] Das S, Forer L, Schonherr S, Sidore C, Locke AE, Kwong A, et al. Next-generation genotype imputation service and methods. Nat Genet. 2016;48:1284–7.

[29] Genomes Project C, Auton A, Brooks LD, Durbin RM, Garrison EP, Kang HM, et al. A global reference for human genetic variation. Nature. 2015;526:68–74.

[30] Zhang D, Dey R, Lee S. Fast and robust ancestry prediction using principal component analysis. Bioinformatics. 2020;36:3439–46.

31. Lambert SA, Wingfield B, Gibson JT, Gil L, Ramachandran S, Yvon F, et al. Enhancing the Polygenic Score Catalog with tools for score calculation and ancestry normalization. Nat Genet. 2024;56:1989–94.

[32] Di Tommaso P, Chatzou M, Floden EW, Barja PP, Palumbo E, Notredame C. Nextflow enables reproducible computational workflows. Nat Biotechnol. 2017;35:316–9.

[33] Ewels PA, Peltzer A, Fillinger S, Patel H, Alneberg J, Wilm A, et al. The nf-core framework for community-curated bioinformatics pipelines. Nat Biotechnol. 2020;38:276–8.

[34] Gruning B, Dale R, Sjodin A, Chapman BA, Rowe J, Tomkins-Tinch CH, et al. Bioconda: sustainable and comprehensive software distribution for the life sciences. Nat Methods. 2018;15:475–6.

[35] Sofer T, Kurniansyah N, Granot-Hershkovitz E, Goodman MO, Tarraf W, Broce I, et al. A polygenic risk score for Alzheimer’s disease constructed using APOE-region variants has stronger association than APOE alleles with mild cognitive impairment in Hispanic/Latino adults in the U.S. Alzheimers Res Ther. 2023;15:146.

[36] Nalls MA, Blauwendraat C, Vallerga CL, Heilbron K, Bandres-Ciga S, Chang D, et al. Identification of novel risk loci, causal insights, and heritable risk for Parkinson’s disease: a meta-analysis of genome-wide association studies. Lancet Neurol. 2019;18:1091–102.

[37] Browning SR, Browning BL. High-resolution detection of identity by descent in unrelated individuals. Am J Hum Genet. 2010;86:526–39.

[38] Andrews S, Krueger F, Segonds-Pichon A, Biggins L, Krueger C, Wingett S. FastQC: a quality control tool for high throughput sequence data. Babraham Institute2012.

[39] Dobin A, Davis CA, Schlesinger F, Drenkow J, Zaleski C, Jha S, et al. STAR: ultrafast universal RNA-seq aligner. Bioinformatics. 2012;29:15–21.

[40] Patro R, Duggal G, Love MI, Irizarry RA, Kingsford C. Salmon provides fast and bias-aware quantification of transcript expression. Nat Methods. 2017;14:417–9.

[41] Wickham H. ggplot2: Elegant Graphics for Data Analysis: Springer-Verlag New York; 2016.

[42] Baden T, Maina MB, Maia Chagas A, Mohammed YG, Auer TO, Silbering A, et al. TReND in Africa: Toward a Truly Global (Neuro)science Community. Neuron. 2020;107:412–6.

[43] Isah MB, Muhammad Z, Lawan MM, Alkhamis AI, Goni BW, Oakley SS, et al. Setting up a state-of-the-art laboratory in resource limited settings: A case study of the biomedical science research and training centre in Northeast Nigeria. European Journal of Neuroscience. 2024;59:1681–95.

[44] Weitkamp E, Larbey R, Maina MB, Petherick K, Muhammad MS, Tsanni A, et al. Science Communication practices and Trust in information sources amongst Nigerian scientists and journalists. Journal of Science Communication. 2023;22:A04.

[45] Kilpinen H, Goncalves A, Leha A, Afzal V, Alasoo K, Ashford S, et al. Common genetic variation drives molecular heterogeneity in human iPSCs. Nature. 2017;546:370–5.

[46] Andrews SJ, Renton AE, Fulton-Howard B, Podlesny-Drabiniok A, Marcora E, Goate AM. The complex genetic architecture of Alzheimer’s disease: novel insights and future directions. EBioMedicine. 2023;90:104511.

[47] Rajabli F, Feliciano BE, Celis K, Hamilton-Nelson KL, Whitehead PL, Adams LD, et al. Ancestral origin of ApoE epsilon4 Alzheimer disease risk in Puerto Rican and African American populations. PLoS Genet. 2018;14:e1007791.

[48] Liu CC, Liu CC, Kanekiyo T, Xu H, Bu G. Apolipoprotein E and Alzheimer disease: risk, mechanisms and therapy. Nat Rev Neurol. 2013;9:106–18.

[49] Leonenko G, Baker E, Stevenson-Hoare J, Sierksma A, Fiers M, Williams J, et al. Identifying individuals with high risk of Alzheimer’s disease using polygenic risk scores. Nat Commun. 2021;12:4506.

[50] Cruchaga C, Del-Aguila JL, Saef B, Black K, Fernandez MV, Budde J, et al. Polygenic risk score of sporadic late-onset Alzheimer’s disease reveals a shared architecture with the familial and early-onset forms. Alzheimers Dement. 2018;14:205–14.

[51] Tan CH, Hyman BT, Tan JJX, Hess CP, Dillon WP, Schellenberg GD, et al. Polygenic hazard scores in preclinical Alzheimer disease. Ann Neurol. 2017;82:484–8.

[52] Hutton M, Lendon CL, Rizzu P, Baker M, Froelich S, Houlden H, et al. Association of missense and 5′-splice-site mutations in tau with the inherited dementia FTDP-17. Nature. 1998;393:702–5.

[53] Dumanchin C, Camuzat A, Campion D, Verpillat P, Hannequin D, Dubois B, et al. Segregation of a Missense Mutation in the Microtubule-Associated Protein Tau Gene with Familial Frontotemporal Dementia and Parkinsonism. Human Molecular Genetics. 1998;7:1825–9.

[54] Saito Y, Geyer A, Sasaki R, Kuzuhara S, Nanba E, Miyasaka T, et al. Early-onset, rapidly progressive familial tauopathy with R406W mutation. Neurology. 2002;58:811–3.

[55] Ghetti B, Oblak AL, Boeve BF, Johnson KA, Dickerson BC, Goedert M. Invited review: Frontotemporal dementia caused by microtubule-associated protein tau gene (MAPT) mutations: a chameleon for neuropathology and neuroimaging. Neuropathol Appl Neurobiol. 2015;41:24–46.

[56] Moore KM, Nicholas J, Grossman M, McMillan CT, Irwin DJ, Massimo L, et al. Age at symptom onset and death and disease duration in genetic frontotemporal dementia: an international retrospective cohort study. The Lancet Neurology. 2020;19:145–56.

[57] Rademakers R, Dermaut B, Peeters K, Cruts M, Heutink P, Goate A, et al. Tau (MAPT) mutation Arg406Trp presenting clinically with Alzheimer disease does not share a common founder in Western Europe. Hum Mutat. 2003;22:409–11.

[58] Lindquist SG, Holm IE, Schwartz M, Law I, Stokholm J, Batbayli M, et al. Alzheimer disease-like clinical phenotype in a family with FTDP-17 caused by a MAPT R406W mutation. Eur J Neurol. 2008;15:377–85.

[59] Ikeuchi T, Kaneko H, Miyashita A, Nozaki H, Kasuga K, Tsukie T, et al. Mutational analysis in early-onset familial dementia in the Japanese population. The role of PSEN1 and MAPT R406W mutations. Dement Geriatr Cogn Disord. 2008;26:43–9.

[60] Consortium GP. A global reference for human genetic variation. Nature. 2015;526:68.

[61] Choudhury A, Aron S, Sengupta D, Hazelhurst S, Ramsay M. African genetic diversity provides novel insights into evolutionary history and local adaptations. Human molecular genetics. 2018;27:R209–R18.

[62] Choudhury A, Aron S, Botigué LR, Sengupta D, Botha G, Bensellak T, et al. High-depth African genomes inform human migration and health. Nature. 2020;586:741–8.

[63] Ramos DM, Skarnes WC, Singleton AB, Cookson MR, Ward ME. Tackling neurodegenerative diseases with genomic engineering: A new stem cell initiative from the NIH. Neuron. 2021;109:1080–3.

[64] Screven LA, Pantazis CB, Andersh KM, Hong S, Vitale D, Lara E, et al. Harnessing diversity to study Alzheimer&#x2019;s disease: A new iPSC resource from the NIH CARD and ADNI. Neuron. 2024;112:694–7.

[65] Fatumo S, Sathan D, Samtal C, Isewon I, Tamuhla T, Soremekun C, et al. Polygenic risk scores for disease risk prediction in Africa: current challenges and future directions. Genome Medicine. 2023;15:87.

[66] Yoo AS, Sun AX, Li L, Shcheglovitov A, Portmann T, Li Y, et al. MicroRNA-mediated conversion of human fibroblasts to neurons. Nature. 2011;476:228–31.

[67] Lu YL, Yoo AS. Mechanistic Insights Into MicroRNA-Induced Neuronal Reprogramming of Human Adult Fibroblasts. Front Neurosci. 2018;12:522.

[68] Capano LS, Sato C, Ficulle E, Yu A, Horie K, Kwon JS, et al. Recapitulation of endogenous 4R tau expression and formation of insoluble tau in directly reprogrammed human neurons. Cell Stem Cell. 2022;29:918–32.e8.

[69] Yang Y, Chen R, Wu X, Zhao Y, Fan Y, Xiao Z, et al. Rapid and Efficient Conversion of Human Fibroblasts into Functional Neurons by Small Molecules. Stem Cell Reports. 2019;13:862–76.

[70] Vierbuchen T, Ostermeier A, Pang ZP, Kokubu Y, Südhof TC, Wernig M. Direct conversion of fibroblasts to functional neurons by defined factors. Nature. 2010;463:1035–41.

[71] Mertens J, Paquola ACM, Ku M, Hatch E, Bohnke L, Ladjevardi S, et al. Directly Reprogrammed Human Neurons Retain Aging-Associated Transcriptomic Signatures and Reveal Age-Related Nucleocytoplasmic Defects. Cell Stem Cell. 2015;17:705–18.

[72] Huh CJ, Zhang B, Victor MB, Dahiya S, Batista LF, Horvath S, et al. Maintenance of age in human neurons generated by microRNA-based neuronal conversion of fibroblasts. Elife. 2016;5.

[73] Victor MB, Richner M, Olsen HE, Lee SW, Monteys AM, Ma C, et al. Striatal neurons directly converted from Huntington’s disease patient fibroblasts recapitulate age-associated disease phenotypes. Nature Neuroscience. 2018;21:341–52.

[74] Minaya MA, Mahali S, Iyer AK, Eteleeb AM, Martinez R, Huang G, et al. Conserved gene signatures shared among MAPT mutations reveal defects in calcium signaling. Frontiers in Molecular Biosciences. 2023;10.

[75] Jiang S, Wen N, Li Z, Dube U, Del Aguila J, Budde J, et al. Integrative system biology analyses of CRISPR-edited iPSC-derived neurons and human brains reveal deficiencies of presynaptic signaling in FTLD and PSP. Transl Psychiatry. 2018;8:265.

[76] Mahali S, Martinez R, King M, Verbeck A, Harari O, Benitez BA, et al. Defective proteostasis in induced pluripotent stem cell models of frontotemporal lobar degeneration. Transl Psychiatry. 2022;12:508.

[77] Bhagat R, Minaya MA, Renganathan A, Mehra M, Marsh J, Martinez R, et al. Long non-coding RNA SNHG8 drives stress granule formation in tauopathies. Mol Psychiatry. 2023;28:4889–901.

