## Supplemental Figures for "Somatic and Stem Cell Bank to Study the Contribution of African Ancestry to Dementia: African iPSC Initiative"

### Supplemental Figure 1

A

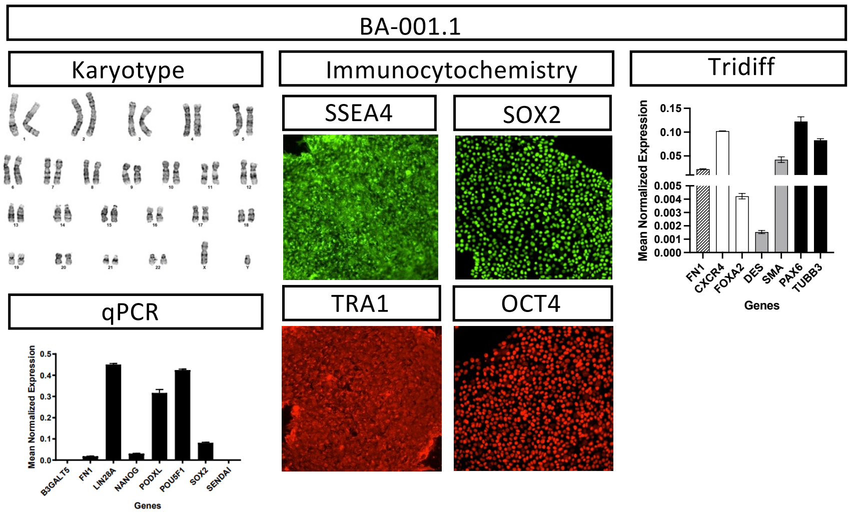


B

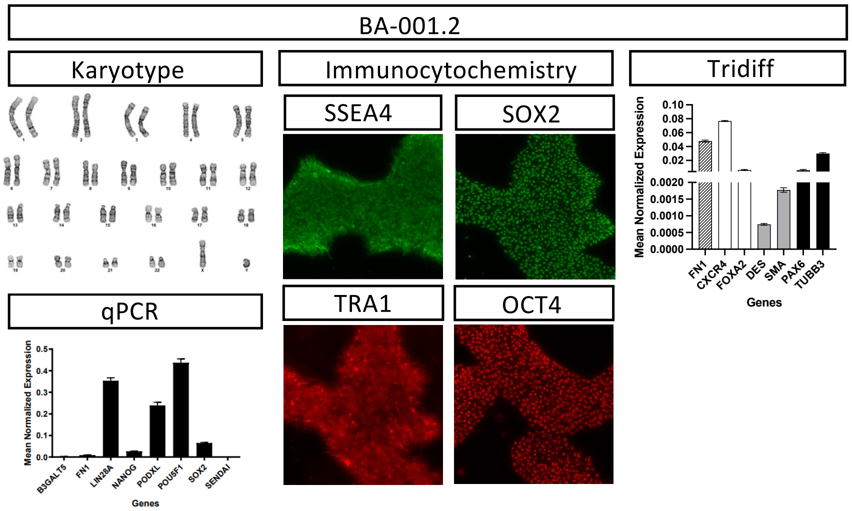


**Supplemental Figure 1: Characterization of iPSC line BA-001.** Representative iPSC characterization data. G-band karyotyping reveals no chromosomal abnormalities. qPCR for pluripotency markers. Immunocytochemistry for pluripotency markers OCT4, SOX2, SSEA4, and TRA-1-60. Differentiation into cells found in each of the three germ layers were validated using qPCR for layer specific markers. A. BA-001.1. B. BA-001.2.

#### Supplemental Figure 2

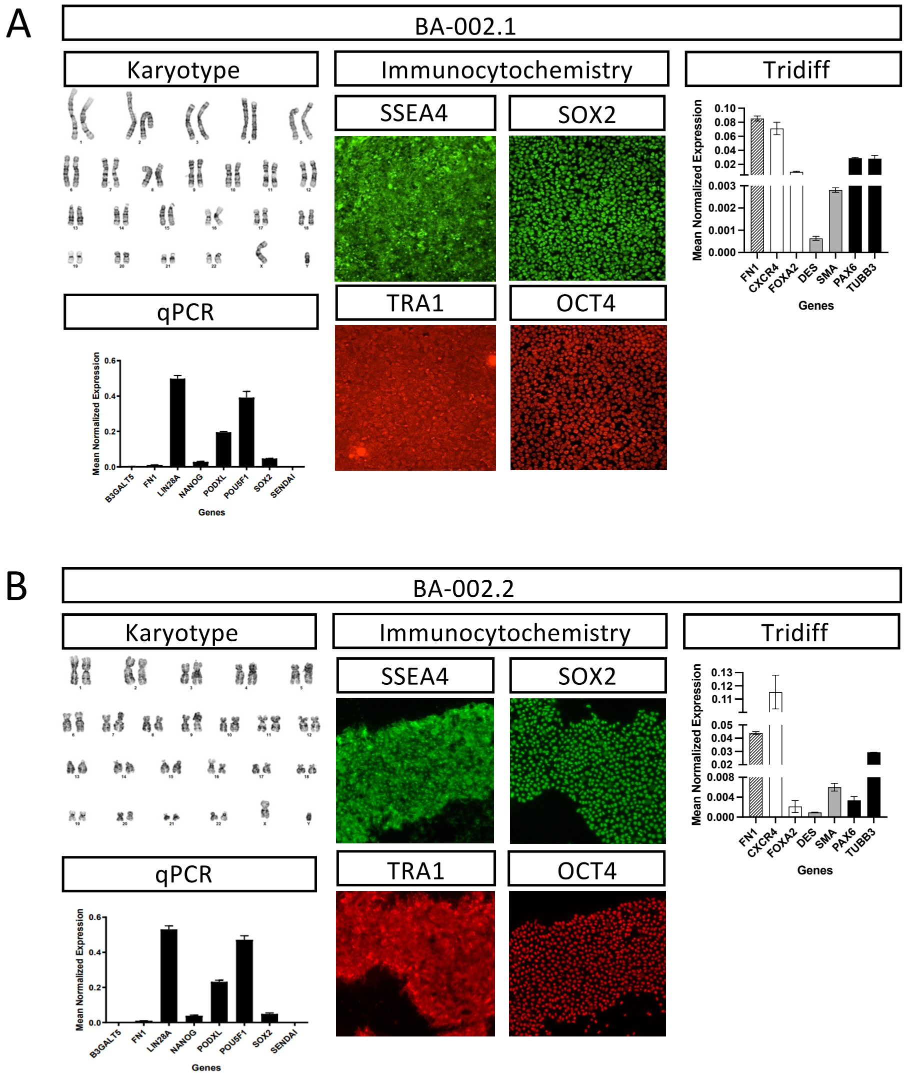


**Supplemental Figure 2: Characterization of iPSC line BA-002.** Representative iPSC characterization data. G-band karyotyping reveals no chromosomal abnormalities. qPCR for pluripotency markers. Immunocytochemistry for pluripotency markers OCT4, SOX2, SSEA4, and TRA-1-60. Differentiation into cells found in each of the three germ layers were validated using qPCR for layer specific markers. A. BA-002.1. B. BA-002.2.

### Supplemental Figure 3

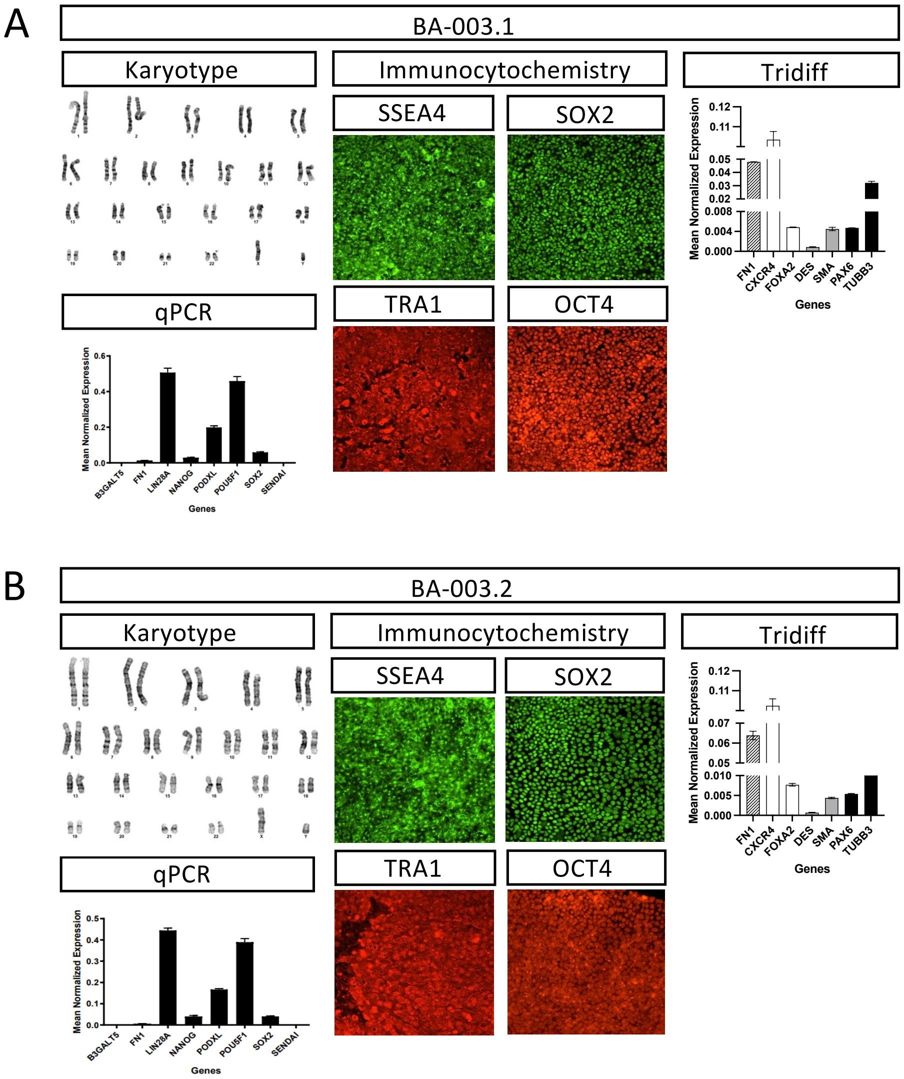


**Supplemental Figure 3: Characterization of iPSC line BA-003.** Representative iPSC characterization data. G-band karyotyping reveals no chromosomal abnormalities. qPCR for pluripotency markers. Immunocytochemistry for pluripotency markers OCT4, SOX2, SSEA4, and TRA-1-60. Differentiation into cells found in each of the three germ layers were validated using qPCR for layer specific markers. A. BA-003.1. B. BA-003.2.

### Supplemental Figure 4

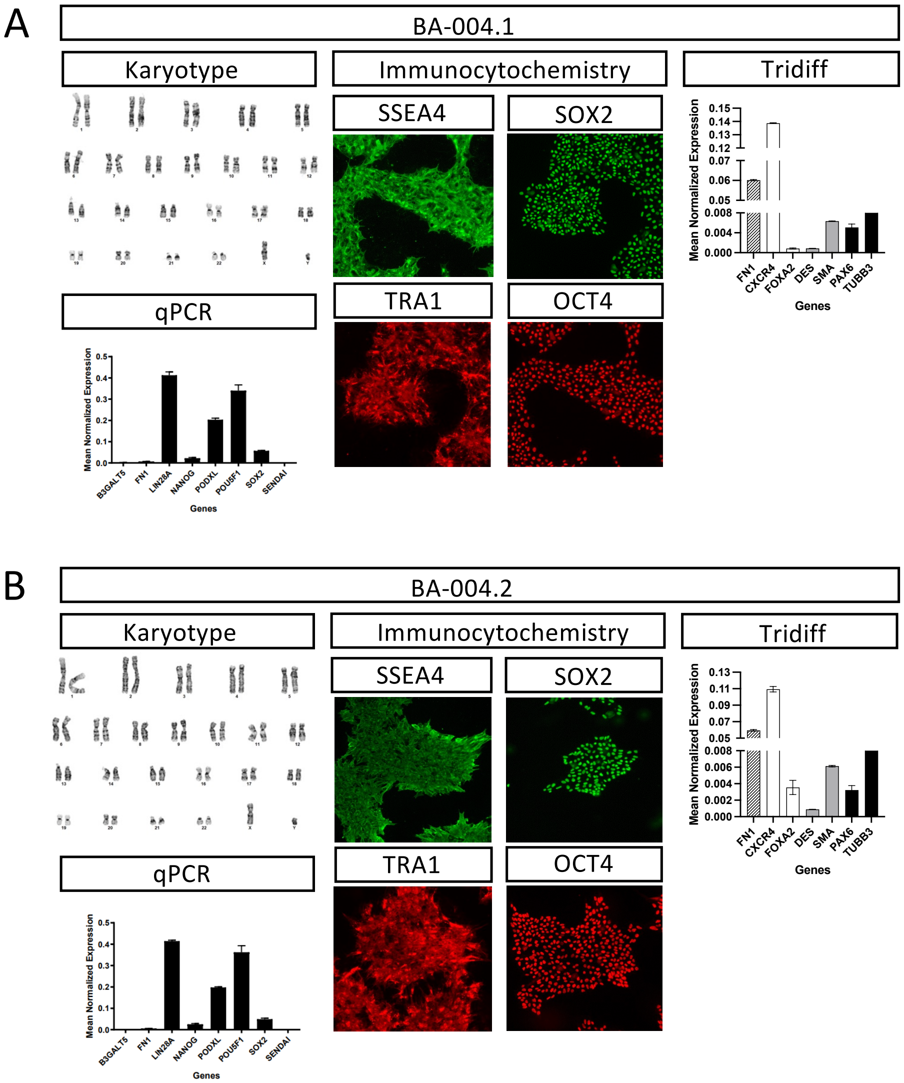


**Supplemental Figure 4: Characterization of iPSC line BA-004.** Representative iPSC characterization data. G-band karyotyping reveals no chromosomal abnormalities. qPCR for pluripotency markers. Immunocytochemistry for pluripotency markers OCT4, SOX2, SSEA4, and TRA-1-60. Differentiation into cells found in each of the three germ layers were validated using qPCR for layer specific markers. A. BA-004.1. B. BA-004.2.

### Supplemental Figure 5

A

B

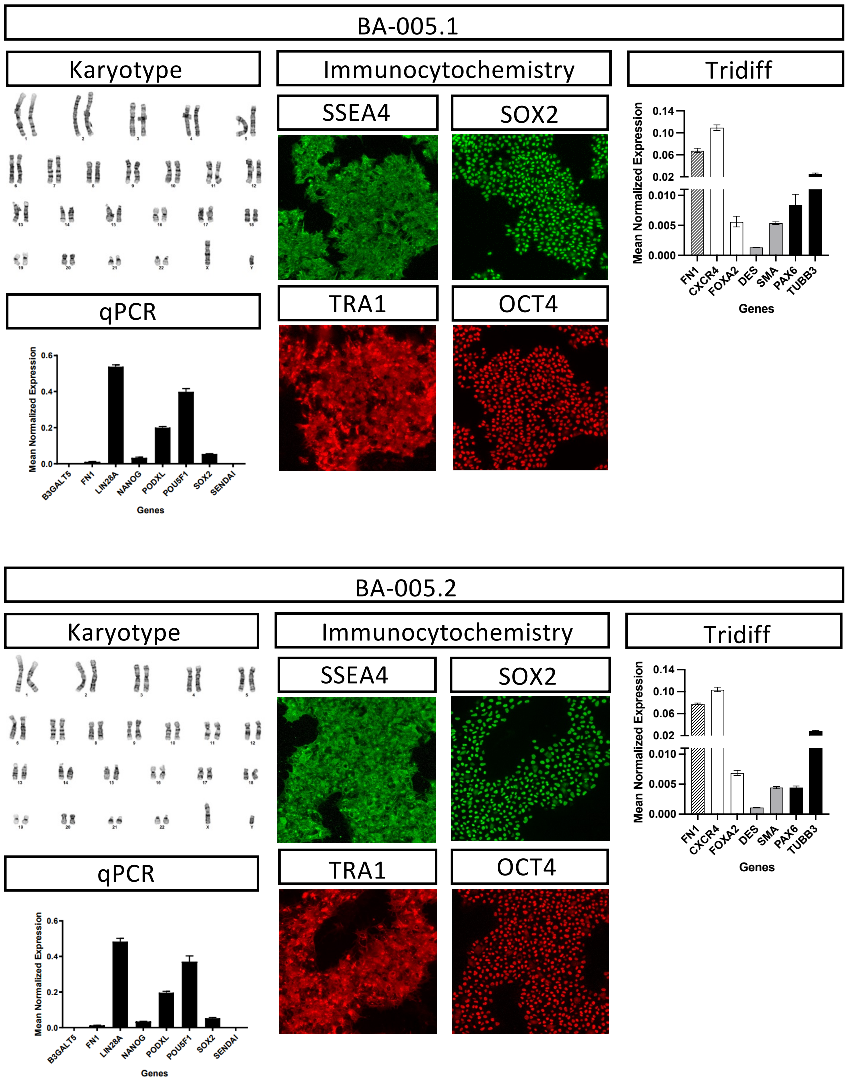


**Supplemental Figure 5: Characterization of iPSC line BA-005.** Representative iPSC characterization data. G-band karyotyping reveals no chromosomal abnormalities. qPCR for pluripotency markers. Immunocytochemistry for pluripotency markers OCT4, SOX2, SSEA4, and TRA-1-60. Differentiation into cells found in each of the three germ layers were validated using qPCR for layer specific markers. A. BA-005.1. B. BA-005.2.

### Supplemental Figure 6

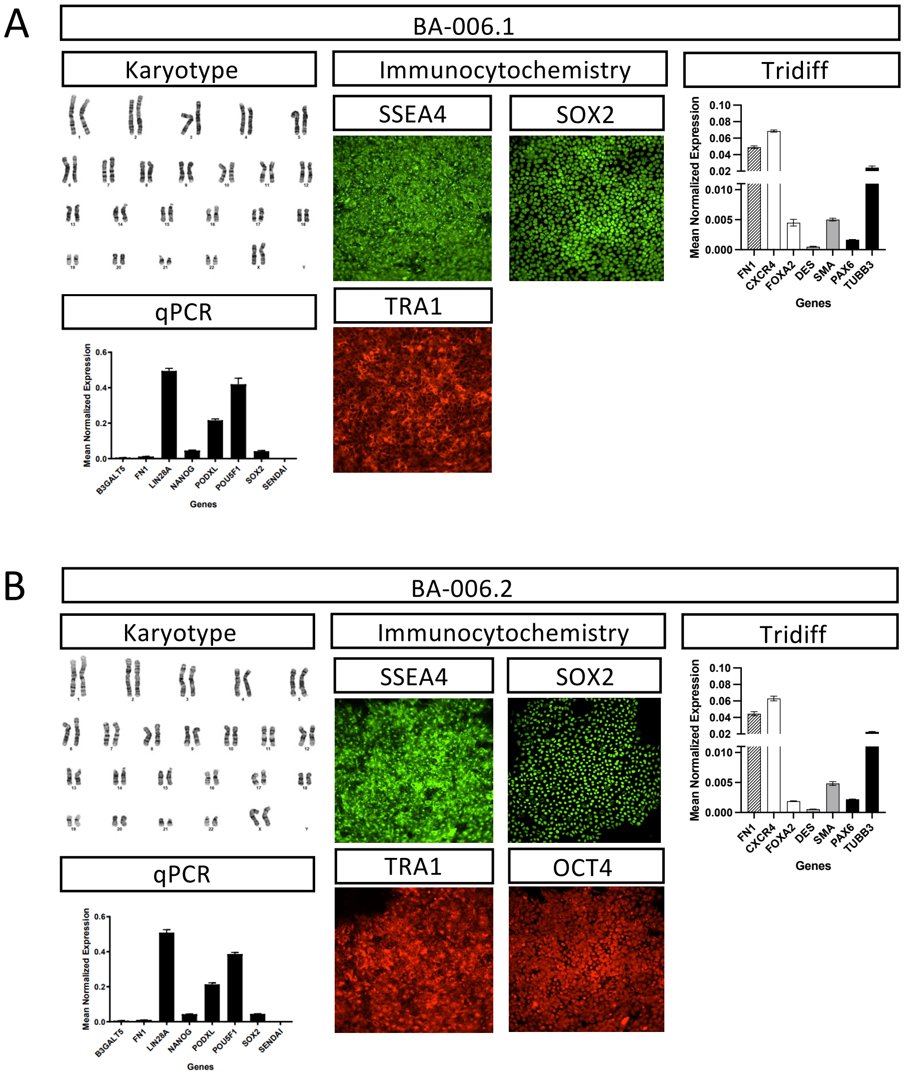


**Supplemental Figure 6: Characterization of iPSC line BA-006.** Representative iPSC characterization data. G-band karyotyping reveals no chromosomal abnormalities. qPCR for pluripotency markers. Immunocytochemistry for pluripotency markers OCT4, SOX2, SSEA4, and TRA-1-60. Differentiation into cells found in each of the three germ layers were validated using qPCR for layer specific markers. A. BA-006.1. B. BA-006.2.

### Supplemental Figure 7

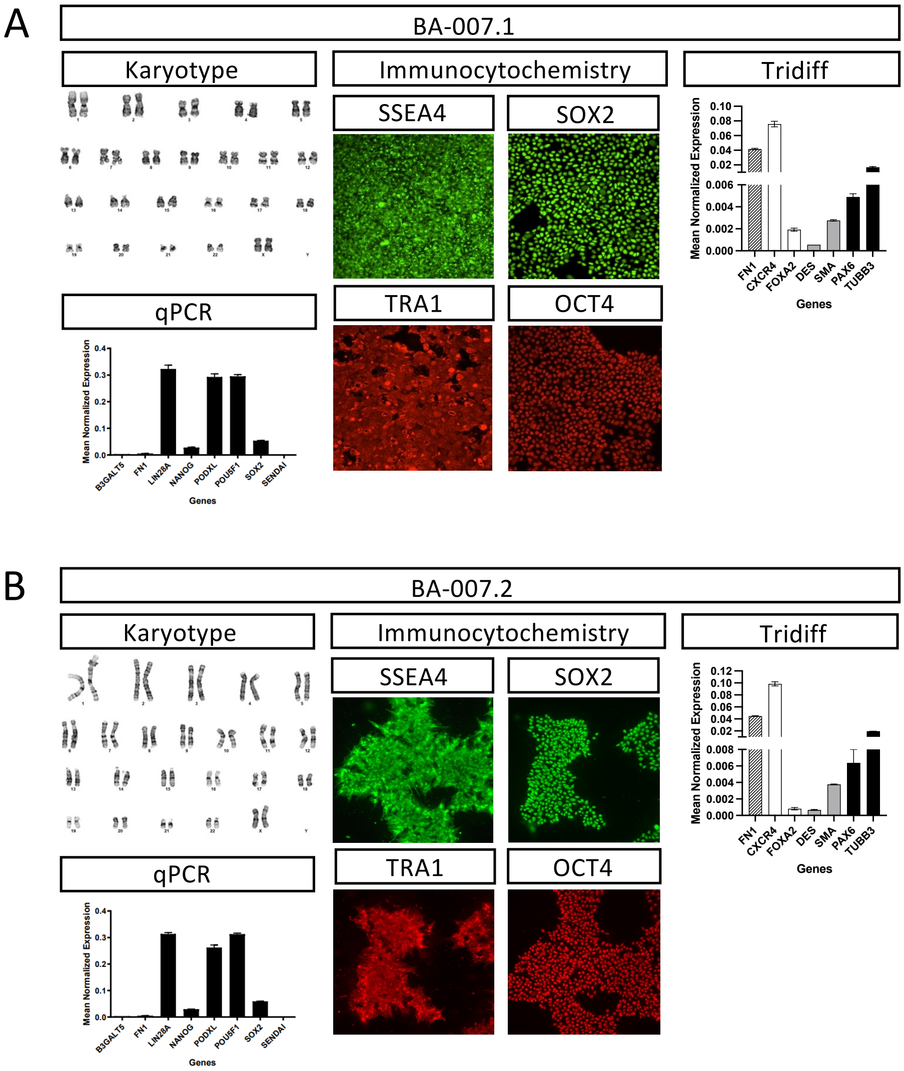


**Supplemental Figure 7: Characterization of iPSC line BA-007.** Representative iPSC characterization data. G-band karyotyping reveals no chromosomal abnormalities. qPCR for pluripotency markers. Immunocytochemistry for pluripotency markers OCT4, SOX2, SSEA4, and TRA-1-60. Differentiation into cells found in each of the three germ layers were validated using qPCR for layer specific markers. A. BA-007.1. B. BA-007.2.

**Supplemental Figure 8**

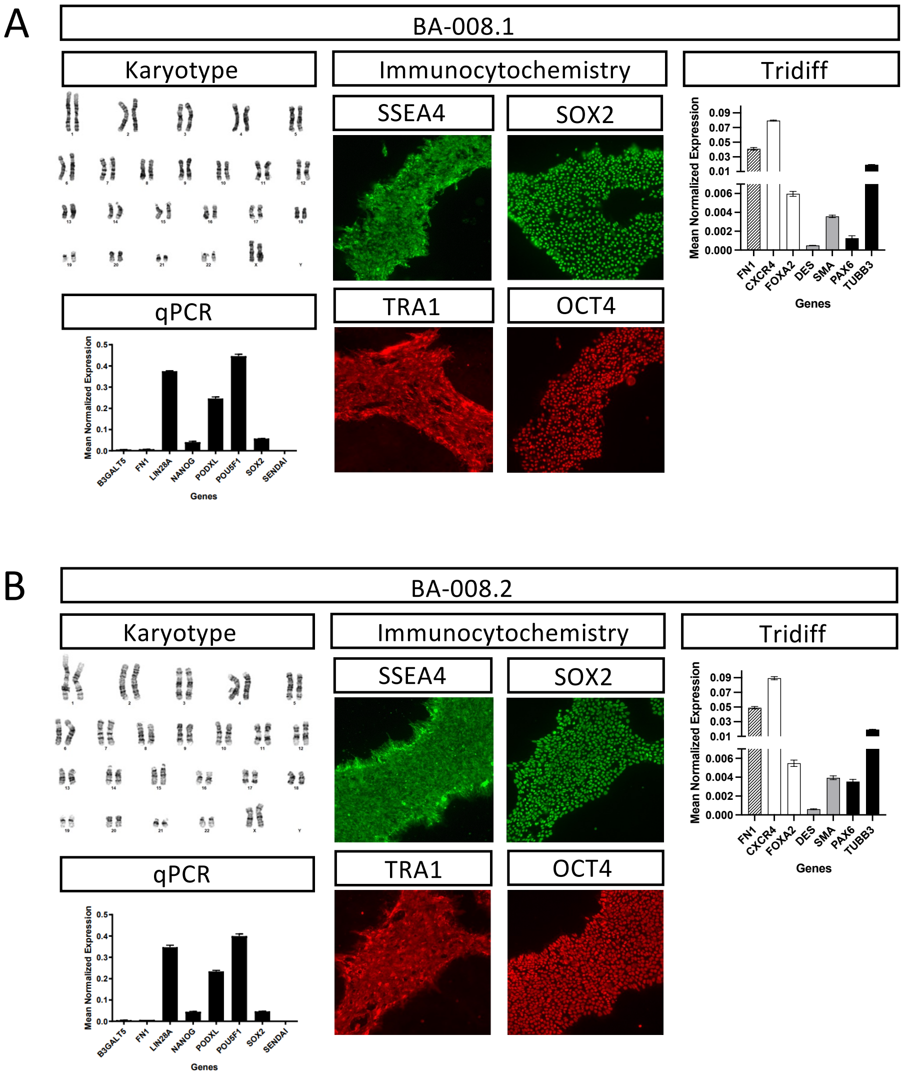


**Supplemental Figure 8: Characterization of iPSC line BA-008.** Representative iPSC characterization data. G-band karyotyping reveals no chromosomal abnormalities. qPCR for pluripotency markers. Immunocytochemistry for pluripotency markers OCT4, SOX2, SSEA4, and TRA-1-60. Differentiation into cells found in each of the three germ layers were validated using qPCR for layer specific markers. A. BA-008.1. B. BA-008.2.

### Supplemental Figure 9

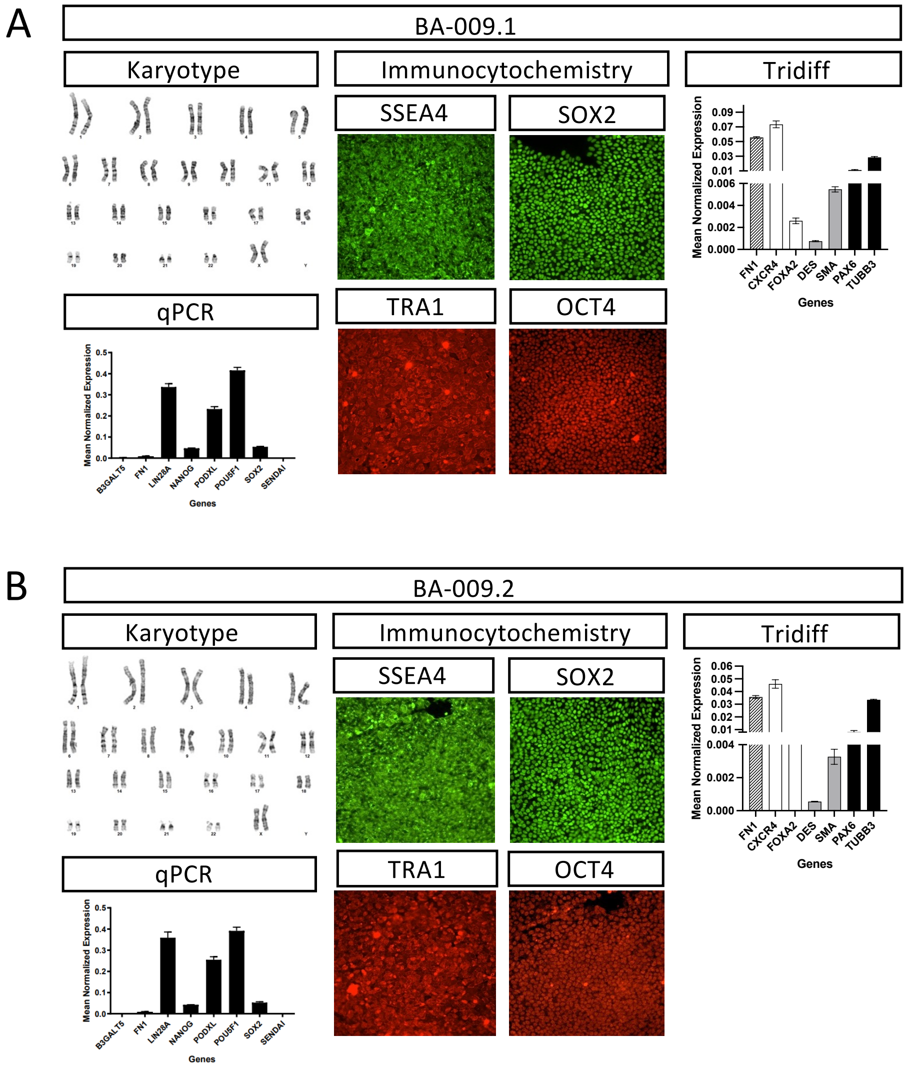


**Supplemental Figure 9: Characterization of iPSC line BA-009.** Representative iPSC characterization data. G-band karyotyping reveals no chromosomal abnormalities. qPCR for pluripotency markers. Immunocytochemistry for pluripotency markers OCT4, SOX2, SSEA4, and TRA-1-60. Differentiation into cells found in each of the three germ layers were validated using qPCR for layer specific markers. A. BA-009.1. B. BA-009.2.

### Supplemental Figure 10

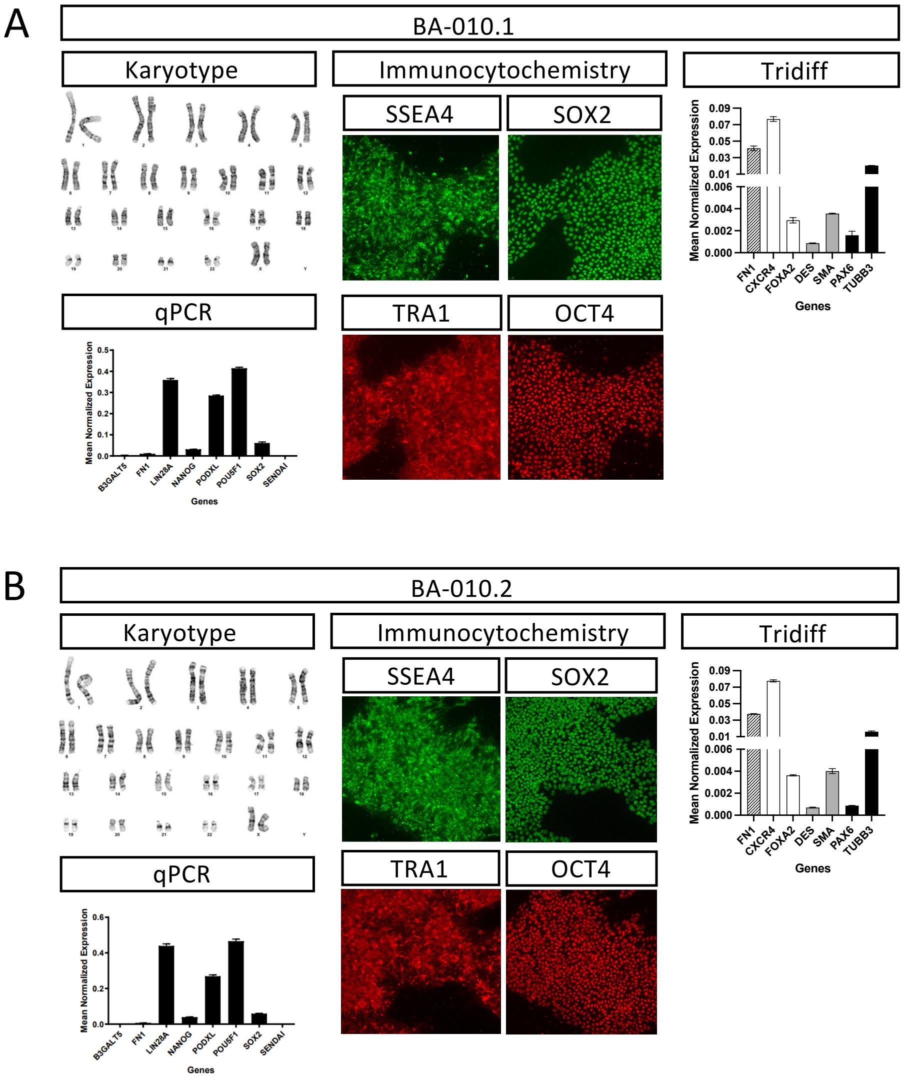


**Supplemental Figure 10: Characterization of iPSC line BA-010.** Representative iPSC characterization data. G-band karyotyping reveals no chromosomal abnormalities. qPCR for pluripotency markers. Immunocytochemistry for pluripotency markers OCT4, SOX2, SSEA4, and TRA-1-60. Differentiation into cells found in each of the three germ layers were validated using qPCR for layer specific markers. A. BA-010.1. B. BA-010.2.

### Supplemental Figure 11

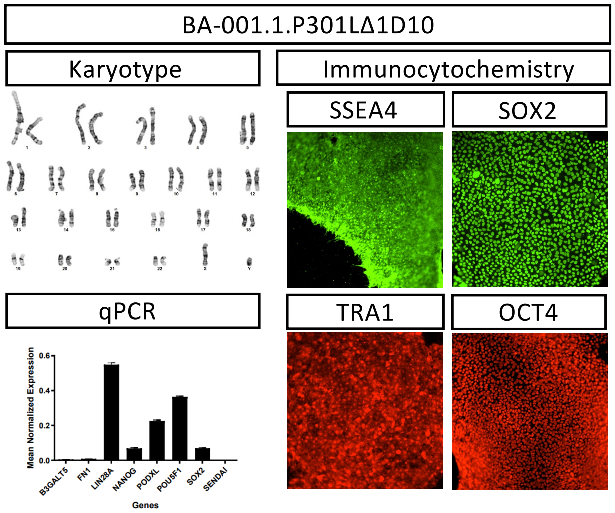


**Supplemental Figure 11: Characterization of iPSC line BA-001.1.P301LΔ1D10.** Representative iPSC characterization data. G-band karyotyping reveals no chromosomal abnormalities. qPCR for pluripotency markers. Immunocytochemistry for pluripotency markers OCT4, SOX2, SSEA4, and TRA-1-60.

### Supplemental Figure 12

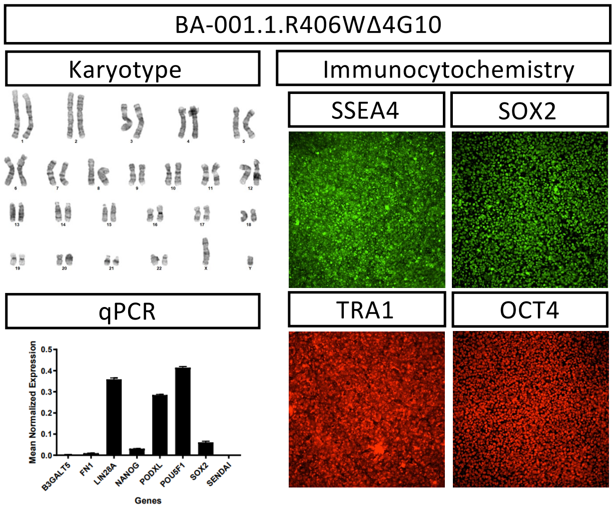


**Supplemental Figure 12: Characterization of iPSC line BA-001.1.R406WΔ4G10.** Representative iPSC characterization data. G-band karyotyping reveals no chromosomal abnormalities. qPCR for pluripotency markers. Immunocytochemistry for pluripotency markers OCT4, SOX2, SSEA4, and TRA-1-60.
